# Cytokine Signatures of Immune Checkpoint Inhibitor Myocarditis and the Modifying Role of Clonal Hematopoiesis: A Machine-Learning and Multi-Omics Study

**DOI:** 10.64898/2026.11.11.26367634

**Authors:** Rachel Jaber Chehayeb, Nancy Manchanda, Dat Hong, Akarvin Jeyashanmugaraja, Jaiveer Singh, James L Cross, Avnith Vijayram, Carlos Matute Martinez, Ritujith Jayakrishnan, Anthos Christofides, Derrick Lin, Yunju Im, Jennifer VanOudenhove, Giulia Biancon, Eiman Ibrahim, Anastasia Ardasheva, Emily Yuan Shuan Lin, Ross Levine, Evaline Cheng, Stephanie Halene, John Hwa, Neal Smith, Chloe Villani, Han Zhu, Alokkumar Jha, Jennifer M Kwan

**Author notes:** co-corresponding authors **Corresponding Author:** Jennifer M Kwan, MD, PhD 300 George St #770b New Haven CT 06511 Twitter: @jennkwanMDPhD. co-first author (Rachel Jaber Chehayeb, Nancy Manchanda).

## Abstract

**Background:** Immune checkpoint inhibitor (ICI) myocarditis affects up to 2% of patients on ICI and has up to a 50% mortality. Clonal Hematopoiesis of Indeterminate Potential (CHIP) is a risk factor for ICI myocarditis.

**Objectives:** To leverage a multi-omics approach to evaluate cytokines associated with ICI myocarditis, immunosuppression resistance, CHIP in ICI myocarditis and implicated pathways and cell type contributions.

**Methods:** 236 cardio-oncology patients were enrolled, including 88 who were on ICI. Cytokine profiling from patients on ICI were sampled and analyzed using machine learning and explainable artificial intelligence. Single-cell multi-omics of peripheral blood from Yale (n=19), Stanford (n=25), and MGH (n=63) evaluated cell-type contributions of cytokines and implicated pathways. Validation of Yale proteomic analyses was done using the Stanford CYTOF dataset.

**Results:** Top features associated with ICI myocarditis included IL13, CXCL10, PDGF-A, CCL13, CCL17 and KITlg. Top KEGG pathways associated with ICI myocarditis and immunosuppression resistant ICI myocarditis included JAK-STAT signaling, IL17 signaling and lipid/atherosclerosis pathways. At the single-cell level, the consensus signals distinguishing myocarditis from non-myocarditis across cohorts was predominantly driven by module-level JAK-STAT, T-cell, and Toll-like receptor activation. Similar transcription gene programs were activated in CHIP carriers including modules implicated in the pro-inflammatory immune response, such as NF-kappa B, IL17, MAPK, and TNF pathways within CHIP cells spanning lymphoid (cytotoxic CD8+ NKT, and natural killer cells) and myeloid lineages.

**Conclusions:** Across cohorts, JAK-STAT signaling emerges as a convergent and potentially actionable axis in ICI myocarditis that is further amplified in CHIP carriers, supporting cytokine-guided risk stratification and JAK-STAT–directed immunosuppression as testable strategies.

## Introduction

While immune check point inhibitors (ICI) have greatly improved survival in several types of cancer, they have also been associated with a variety of immune-related adverse events (irAE). Of all irAEs, ICI-associated myocarditis, while rare at 1-2%, is the most fatal with a large proportion of patients suffering from major adverse cardiac events (MACE) and a reported mortality rate of up to 50% (1). Up to 66% of patients can have immunosuppression resistant ICI myocarditis (2).

Our current understanding of the mechanism of PD-1 and PD-L1 inhibitor mediated toxicity involves activation of CD4+ T cells and macrophages resulting in infiltration of the myocardium and activation of the inflammatory NF-κB pathway and single cell studies have identified a unique cell population that was associated with CCL4, CCL4L2, and CCL5 expression in patients with ICI myocarditis (3–5).

Despite these insights, clinical risk stratification for ICI myocarditis remains limited. Prior studies have identified dual ICI therapy, ICI combined with other anti-cancer treatments, female sex, age greater than 75, diabetes mellitus, and cardiovascular disease at baseline as risk factors for ICI myocarditis, but the most consistent risk factor is the use of dual ICI (6). More recently, our team identified a novel risk factor for ICI myocarditis, clonal hematopoiesis of indeterminate potential (CHIP). CHIP is associated with a 2-fold increased risk of ICI myocarditis as well as cardiomyopathy during cancer treatment (7–9). CHIP is a proinflammatory condition characterized by the presence of a clonal, pre-malignant population of hematopoietic cells and cell lineages that harbor a leukemogenic mutation. Myeloid lineage CHIP has been shown to be an independent risk factor for adverse cardiac events and all-cause mortality, and to worsen cancer outcomes and shorten survival (10).

Improved identification of patients at highest risk for ICI-associated myocarditis and immunosuppression-resistant disease is needed to guide cardiac surveillance and inform immunosuppressive strategies. To address these needs, we conducted a prospective cardio-oncology cohort study at Yale University to identify cytokines in peripheral blood that are associated with and predictive of ICI myocarditis and immunosuppression resistant myocarditis. Additionally, given that CHIP has been shown to induce a pro-inflammatory state and be an independent risk factor for ICI-myocarditis, we aimed to assess whether the total number of CHIP variants and particular CHIP variants were associated with higher rates of ICI myocarditis. Additionally, we evaluated the CHIP risk score of our cardio-oncology cohort and cytokine features associated with the intermediate and high-risk groups (11,12). Finally, to further validate identified enriched pathways, characterize cell type contributions of cytokines in ICI myocarditis broadly, as well as in patients with CHIP who develop ICI myocarditis specifically, multi-omics (single cell RNA sequencing and proteomic (cyTOF)) analyses was performed on peripheral blood mononuclear cell tissue samples using both internal and external data. We hypothesized that ICI myocarditis is driven by identifiable peripheral cytokine programs converging on JAK-STAT and allied pro-inflammatory signaling, and that clonal hematopoiesis amplifies these programs at the single-cell level — together providing a rationale for cytokine-guided risk stratification and pathway-directed immunosuppression.

## Methods

### Study Design

236 cardio-oncology patients with solid tumors receiving treatment with chemotherapy and/or immunotherapy at Yale New-Haven Hospital were enrolled in the study. Additionally, twelve patients with breast cancer from MSKCC and 6 non-cancer controls were also included to increase our control numbers for cytokine profiling machine learning training dataset. Patients with any active hematologic malignancy were excluded. The study was approved by the Yale Institutional Review Board and participants provided consent. Patients were enrolled between Jan 1, 2018, and Dec 31, 2023.

### Data Collection

#### Assessing CHIP status and Cytokine profiling

All patients received a blood draw at time of enrollment to assess for CHIP status and for cytokine profiling. Details on patient demographics, baseline comorbidities and outcomes data collection are provided in **Supplemental Methods Text 1**.

To determine CHIP status, we performed whole exome sequencing using a dedicated gene panel against known CHIP genes (13,14), achieving a median of 1000x sequencing depth, as has been previously described (15). Details on the CHIP variant annotation and cytokine profiling are provided in **Supplemental Methods Text 2**.

### Single-cell CHIP inference (CIPHER-CHIP)

To resolve CHIP-associated immune clones from single-cell transcriptomes, we developed a clone-aware probabilistic framework, CIPHER-CHIP (Clone-Informed Probabilistic Hematopoietic Expression Resolution for CHIP). Details on the CIPHER-CHIP pipeline are provided in **Supplemental Methods Text 3**.

### Machine Learning Model

To assess which cytokines were significantly associated with and predictive of our outcomes of interest, we computed point-biserial correlation coefficients and built predictive machine learning models respectively using Python packages (including numpy 1.26.2, scikit-learn 1.3.0). Since steroids alter cytokine testing results, we excluded patients (n=35) with myocarditis who had already initiated steroids from the machine learning model specifically. For our predictive model, we opted for XGBoost. Details of model validation and SHAP-based interpretation of feature directionality are provided in **Supplemental Methods Text 4.**

KEGG pathway enrichment analysis of cytokines identified through both machine learning models and point-biserial correlation were performed through the Enrichr website (https://maayanlab.cloud/Enrichr/) on March 12^th^, 2024 and April 17^th^, 2024 (16). All analysis was performed in Python 3.9.18.

### Multi-omics (scRNA-seq and cyTOF) and pathway enrichment analysis

For pathway enrichment analysis, we used RNA seq data from peripheral blood mono-nuclear cell tissue samples (PBMC) collected from patients after exposure to ICI from the Yale, MGH and Stanford cohorts for the single-cell pathway (AUCell and VISION) analyses, with Stanford single-cell and CyTOF data contributing to cytokine and multi-omics validation, (5,17). Details on the composition of the three cohorts are provided in **Supplemental Methods Text 5**. **Supplemental Methods Text 6** details the Yale single-cell RNA-seq library construction and sequencing protocol. Methods on single cell RNA-seq data integration and cell clustering for the Yale, MGH, and Stanford cohort at provided in **Supplemental Methods Text 7.** Methods on pathway enrichment characterization are provided **in Supplemental Methods Text 8** and **Supplemental Table 6.**

### Multi-Omics Factor Analysis (MOFA)

**Supplemental Text 9** details MOFA methodology including CyTOF data processing for the Stanford cohort.

## Results

### Patients with cancer have a high prevalence of CHIP and patients with CHIP developed ICI myocarditis at a higher rate

We enrolled 236 patients, most of whom had an inpatient consult or outpatient referral to cardiology or cardio oncology (**Table 1**). Of the 236 patients, 88 (37.29%) were receiving ICI therapy, of whom 50 (56.8%) developed ICI myocarditis. Of the 50 patients, 24 (48.0%) developed immunosuppression resistant (IR) ICI myocarditis. Mean age of all patients was 64 with a standard deviation of 13 years. CHIP was highly prevalent in our cohort, with 90 patients (38.1%) harboring CHIP. The rate of CHIP was higher among patients who developed ICI myocarditis than among those who did not (64% [32/50] vs 26% [10/38]). The following were the most common cancer types: breast (41.9% (99/236), genitourinary (14.4% (34/236), lung (15.7% (37/236)), and melanoma (5.9% (14/236) (**Table 2**). Among all patients, 35.2% (83/236) had metastatic disease. Comorbidities, prior oncologic therapy and cardiovascular medications are also listed in **Table 1**. Among all patients, 28% (66/236) developed cardiomyopathy during the study, 21.2% (50/236) developed diastolic dysfunction, and 23.7% died in our monitoring period (56/236). The cardiomyopathy and mortality rates were highest among patients who developed ICI myocarditis (70.0% (35/50) and 60% (30/50) respectively). 216 patients (91.52%) had a baseline echo or cardiac MRI, and 177 (75.0%) had follow up echo or cardiac MRI (**Table 1**). Of patients with ICI myocarditis, 35 had received steroids at time of blood draw and were excluded from the subsequent machine learning analysis.

### Patients with CHIP on ICI who developed myocarditis had higher rates of TET2 mutations and lower rates of IDH2 mutations

In evaluating CHIP gene distribution amongst patients, the most common CHIP gene variants included DNMT3A (35.9%), TET2 (14.1%), PPM1D (10.9%), EZH2 (10.9%), TP53 (8.7%) and IDH2 (7.6%). Of those with CHIP, 10% of the patients had high risk genes such as SF3B1, SRSF2, ZRSR2, JAK2, TP53, RUNX1, FLT3, IDH1 and IDH2, 9% had a VAF >10%, 6% had a VAF of >20% and 14% had ≥2 CHIP variants (**Figure 1a**). The distribution of the total number of CHIP mutations per patient stratified by myocarditis outcome is noted in **Supplemental Table 7** and **Supplemental Figure 1**, with patients with myocarditis having increased CHIP mutation burden. **Figure 1b** and **Supplemental Table 8** demonstrates CHIP gene variant frequencies stratified by myocarditis status.

**Figure 1.**
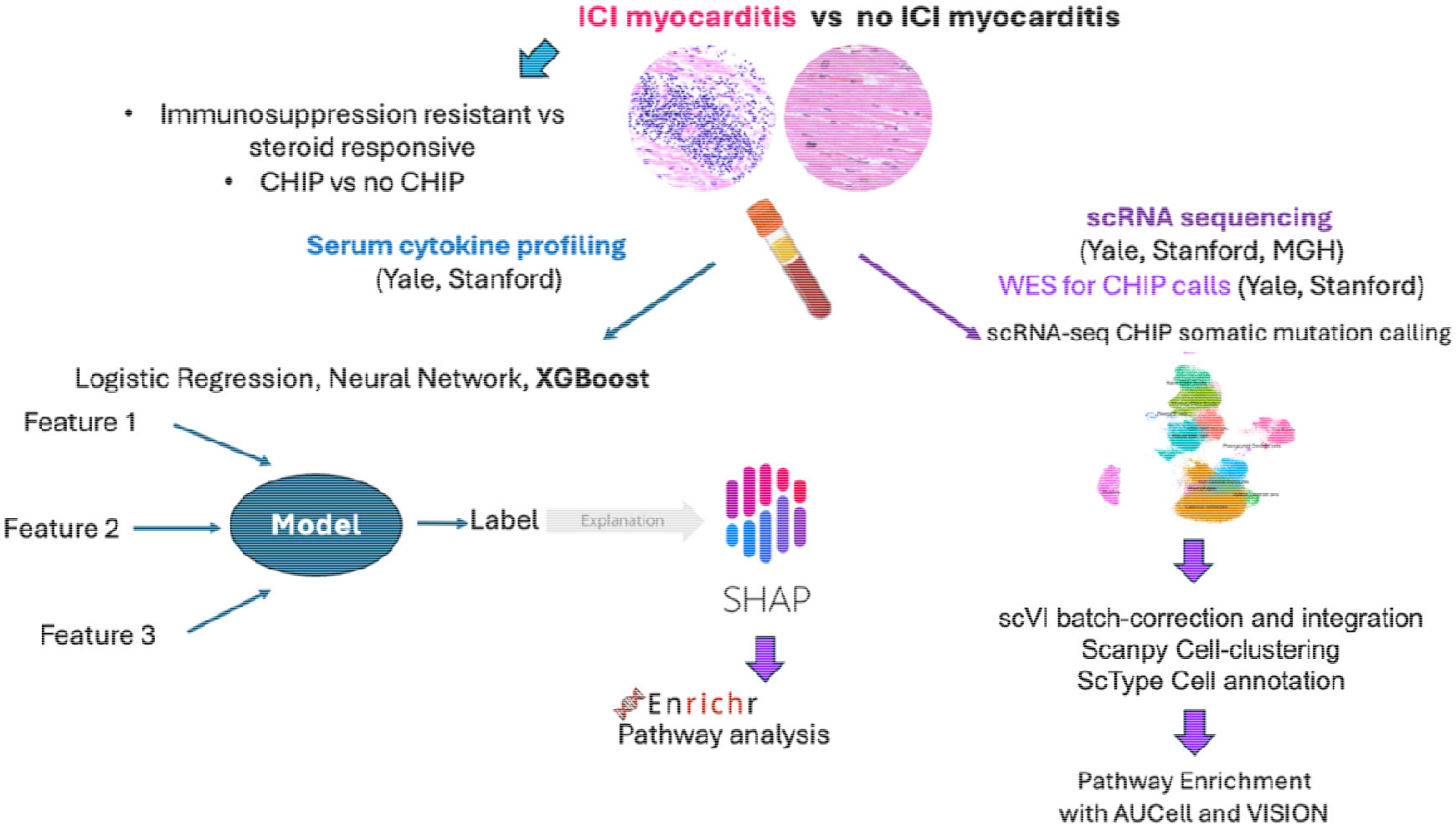
**a:** CHIP gene variant frequency (%) among entire cohort Figure 1b: Bar Graphs of CHIP Variant Frequencies among CHIP Patients on ICI with myocarditis (top) and among CHIP Patients on ICI who did not develop myocarditis (bottom)

While not statistically significant, CHIP patients on ICI who developed myocarditis had higher rates of TET2 mutations (22.9% vs 10%, p=0.66) and lower rates of IDH2 mutations (0% vs 10%, p=0.22) compared to CHIP patients on ICI who did not develop myocarditis. Among patients with ICI, there was no statistically significant difference in cancer subtypes stratified by CHIP status (**Supplemental Table 9**). Demographic and clinical data for the 12 breast cancer patients from MSKCC and the 6 non-cancer controls are shown in **Supplemental Table 10.**

### Four cytokines, PDGF-A, VEGF-A, BCA-1/CXCL13 and TRAIL/TNFSF10, were significantly associated with myocarditis

By point biserial correlation, four cytokines, PDGF-A, VEGF-A, BCA-1/CXCL13 and TRAIL/TNFSF10 were significantly associated with myocarditis (**Supplemental Figure 2**). Additionally, BCA-1/CXCL13, TRAIL/TNFSF10, MIG/CXCL9 and GroA/CXCL1 were significantly associated with the immunosuppression resistant phenotype, as seen in the corresponding correlation plots (**Supplemental Figure 3**).

### IP10/CXCL10, TRAIL/TNFSF10, and MCP-1/CCL2, among other cytokines, are predictive of ICI myocarditis

Among various models including logistic regression, random forest, and multi-layer perceptron, optimized XGboost was the best performing model and identified cytokines predictive of ICI myocarditis (AUC of 0.914, Accuracy of 0.904, F1 score of 0.732) and immunosuppression resistant ICI myocarditis (AUC of 0.826, Accuracy of 0.789, F1 score of 0.724).

Top cytokine features predictive of ICI myocarditis were IL13, IP10/CXCL10, PDGF-A, TRAIL/TNFSF10, IL5, MCP-1/CCL2 and MCP-4/CCL13, TARC/CCL17, SCF/KITLG and FGF2 (Figure 2). **Supplemental Table 11** shows gene names corresponding to identified cytokines. The SHAP model revealed the directionality of the top performing features (**Figure 2**). Cytokines positively associated with ICI myocarditis included IP-10/CXCL10, PDGF-A, MCP-4/CCL13, TARC/CCL17 and SCF/KITLG, whereas IL-13, TRAIL/TNFSF10, IL-5, MCP-1/CCL2 and FGF-2 were negatively associated (**Figure 2**).

**Figure 2:**
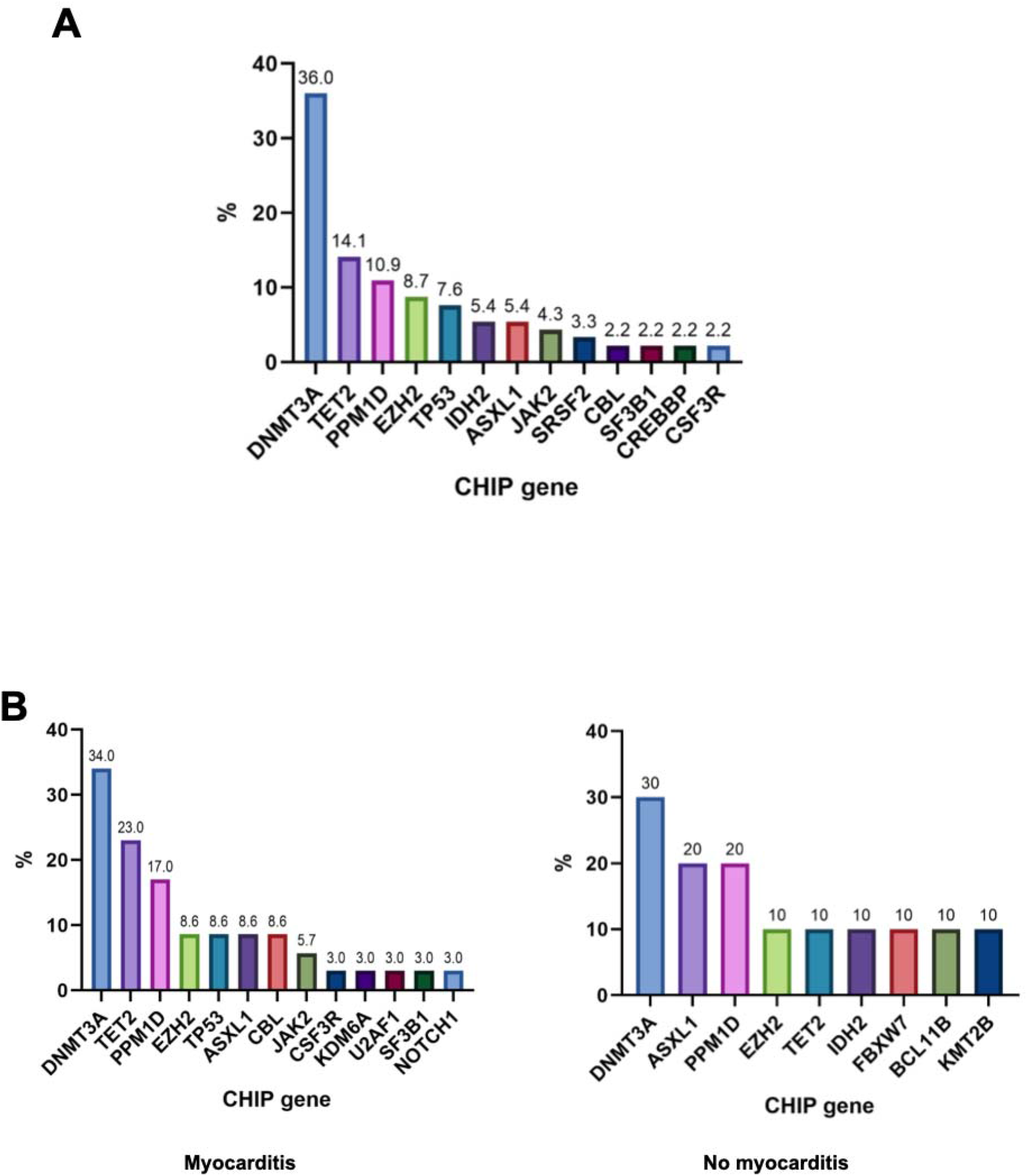
SHAP model (top) and XGboost machine learning (bottom) for top cytokine features associated with ICI myocarditis. The mean absolute value of the SHAP value show in Figure 2 (top) shows the relative strength of the upregulation or downregulation of identified cytokines in terms of impact on the outcome of ICI myocarditis in order of decreasing strength, with variations in IL13 having the greatest predictive value. In the bottom graph, each dot represents a cytokine from one patient sample, with the pink to blue scale representing highest to lowest feature value in predicting the outcome of ICI myocarditis in a particular patient case. For IL13, the clustering of high feature values associated with negative SHAP values implies that downregulation of IL13 is predictive of development of ICI myocarditis.

### JAK-STAT pathway is implicated in ICI myocarditis pathogenesis

KEGG pathway analysis identified 42 pathways with significant association with ICI myocarditis, including the JAK-STAT, IL17, and TNF alpha pathways (**Figure 3, Supplemental Table 12**). The main cytokines associated with the JAK-STAT pathways were IL (interleukin) 5, IL15, IL13, PDGF (platelet-derived growth factor) A, and IL12B, whereas the main cytokines associated with IL17 signaling were CXCL10, IL5, IL13, CCL2, CCL17 (**Table 3**).

**Figure 3:**
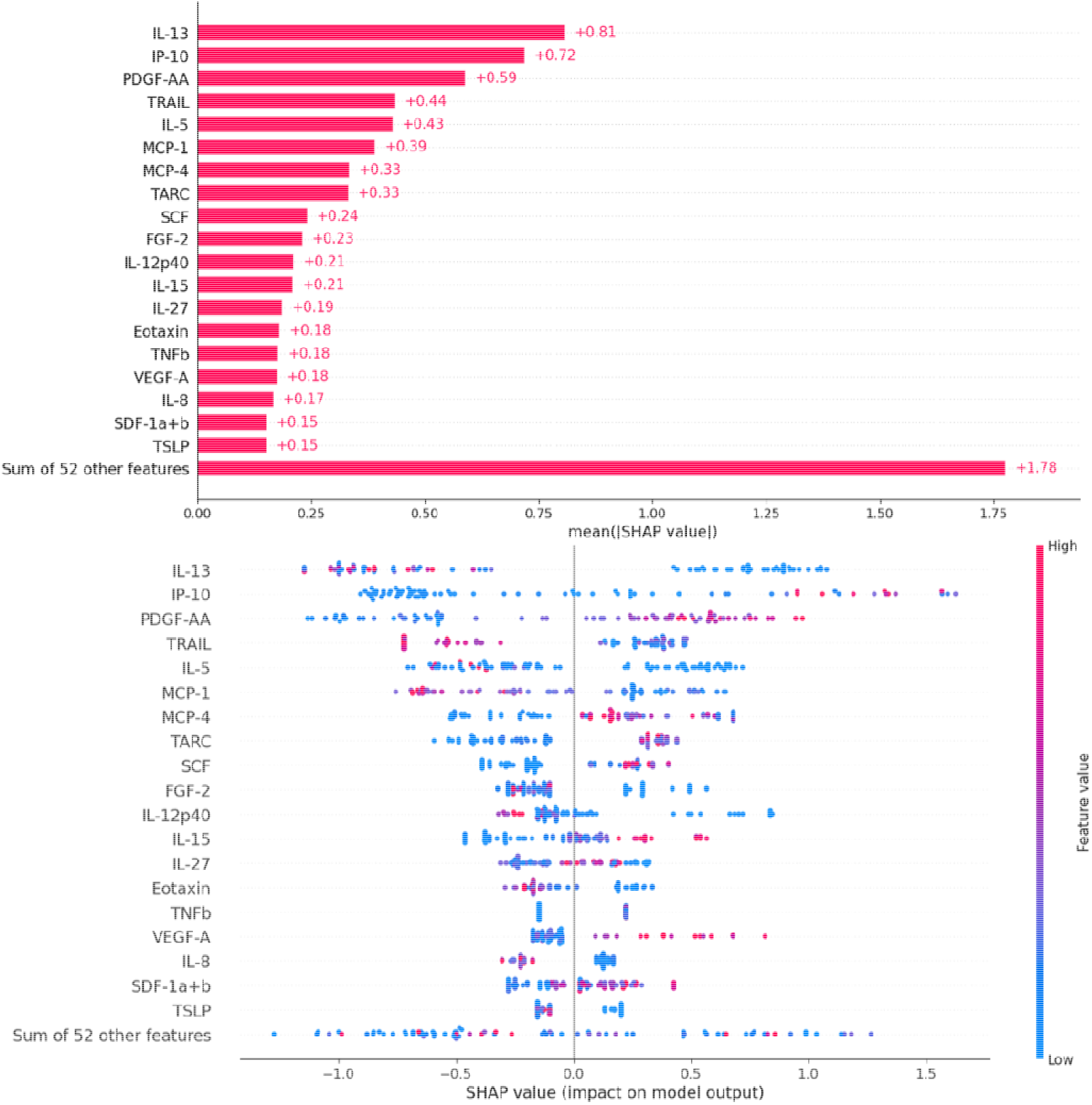
UMAP scatterplot of top KEGG pathways implicated in ICI myocarditis The UMAP scatterplot on the left clusters similar gene sets together, with points outlined in black representing significantly enriched gene sets. This plot is generated by enrichr using the Leiden algorithm with points plotted on the first two UMAP dimensions (16). Highlighted pathways on the right indicate pathways of particular interest given previous associations with immunotherapy mechanisms and toxicity as elaborated on in the discussion.

### JAK-STAT pathway is also implicated in immunosuppression resistant ICI myocarditis

For immunosuppression resistant myocarditis, **Figure 4** shows top predictive cytokines. Cytokines positively associated with the immunosuppression-resistant phenotype included MIP-1b/CCL4, IL-10, Eotaxin-2/CCL24, ENA-78/CXCL5 and BCA-1/CXCL13, whereas MDC/CCL22, TNF-b, IL-1A, FGF-2, EGF and GRO-a/CXCL1 were negatively associated (**Figure 4**). Top KEGG pathways implicated by the cytokines include JAK-STAT, IL17, NFKB, MAPK (**Figure 5**) and the associated pathways and cytokines are featured in **Table 4**. In total, cytokines associated with 44 KEGG pathways were significantly enriched in immunosuppression resistant myocarditis (**Supplemental Table 13**).

**Figure 4:**
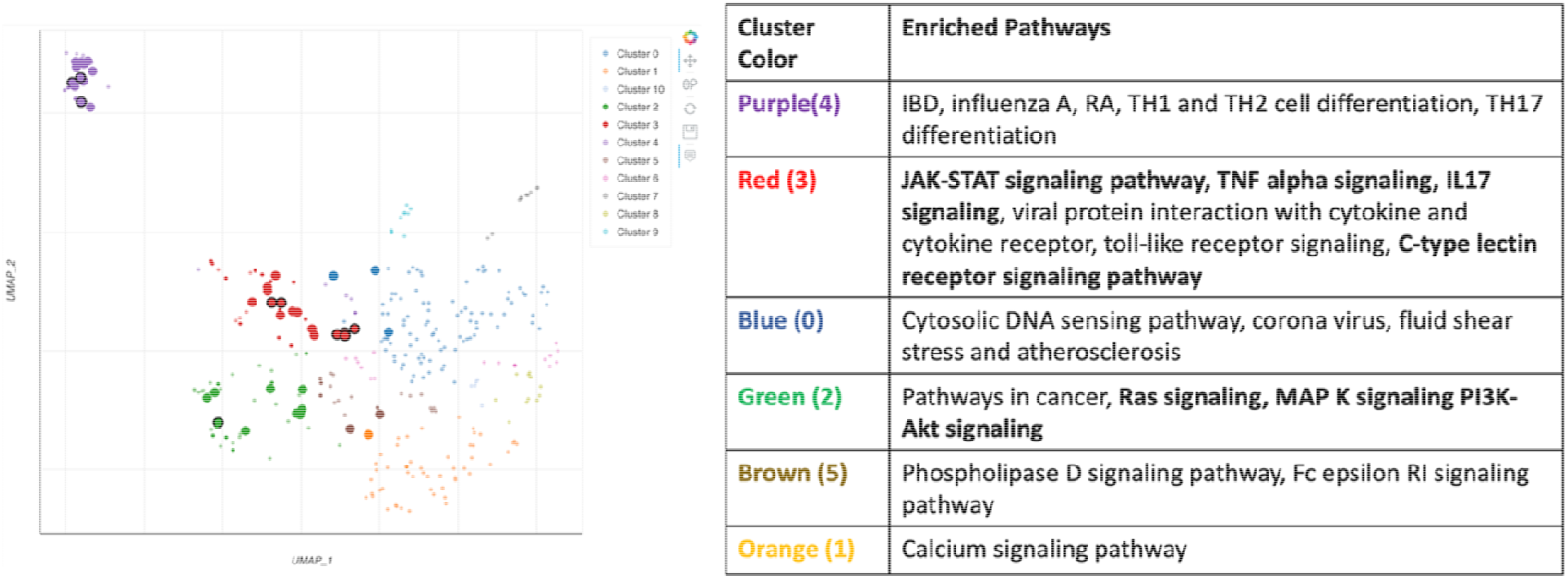
SHAP model (top) and XGboost machine learning (bottom) for top cytokine features associated with immunosuppression resistant ICI myocarditis

**Figure 5:**
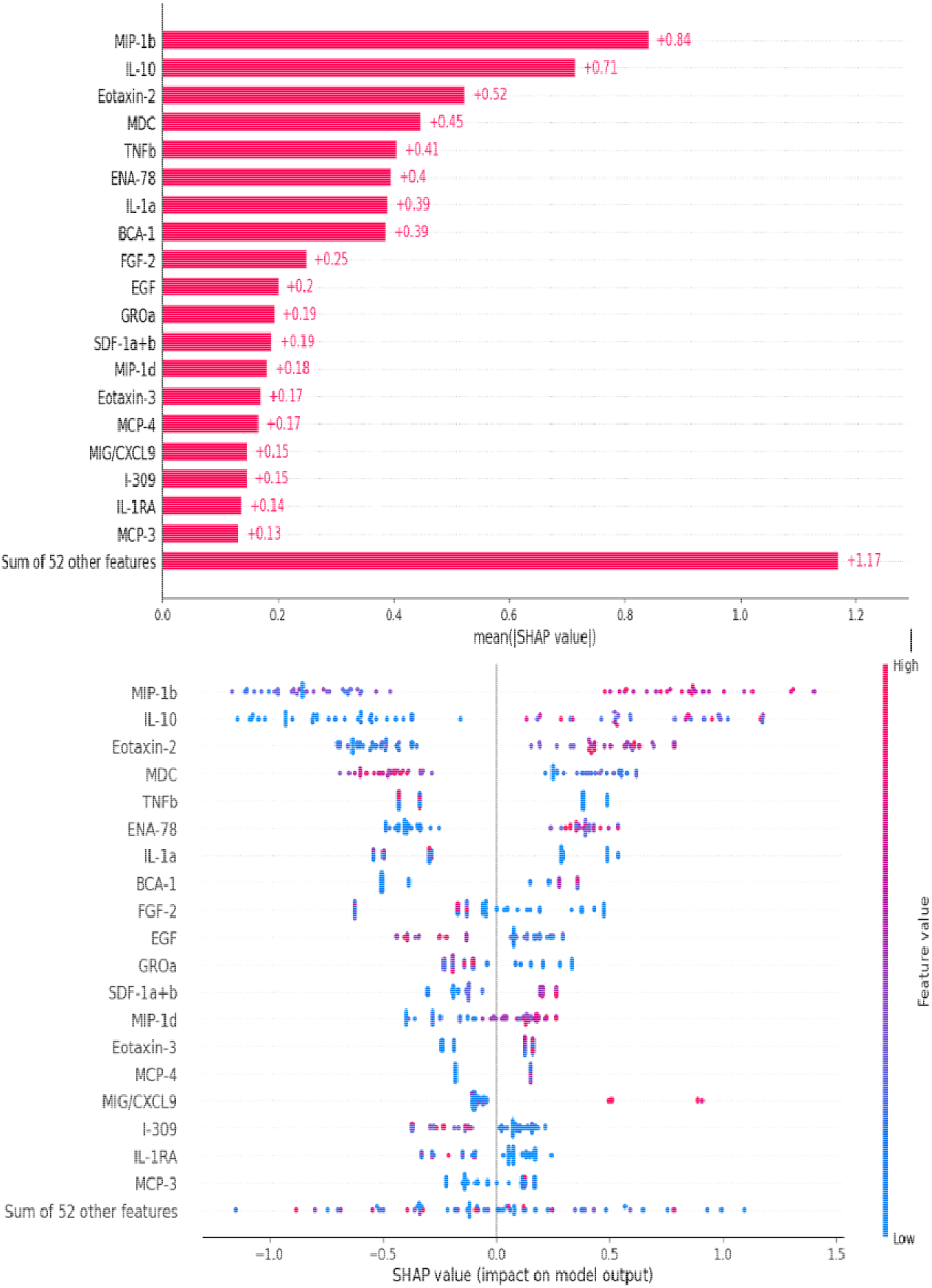
UMAP plot of top KEGG pathways implicated in immunosuppression resistant ICI myocarditis

### JAK inhibition may be beneficial in immunosuppression resistant ICI myocarditis patients

We followed troponin trends for two patients with immunosuppression resistant ICI myocarditis and noted significant improvement in their troponin levels after introducing a JAK inhibitor (JAKi), as shown in **Figure 6**.

**Figure 6:**
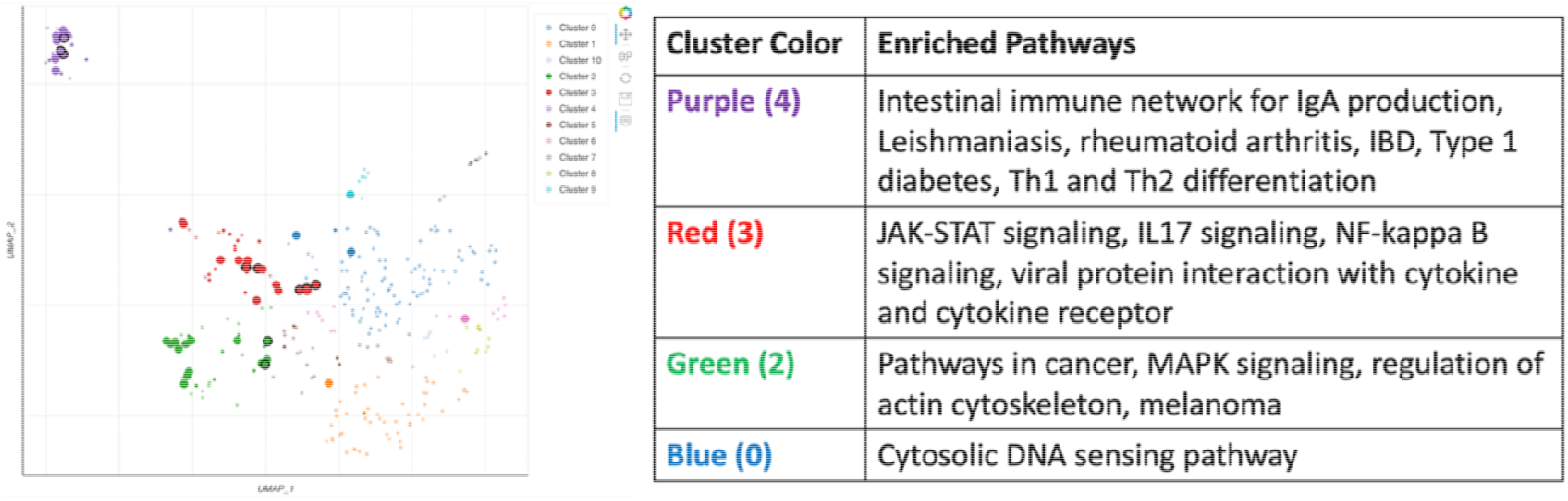
Troponin trend for immunosuppression resistant ICI myocarditis patient 1 (A) and patient 2 (B) and response to JAK inhibitor. X-axis represents time with timing and duration of various immunosuppressive therapies received highlighted.

Patient 1 had a history of endometrial cancer and had been started on pembrolizumab 1 month prior presented with significantly elevated high sensitivity troponins in the 600 ug/L range and a new right bundle branch block (**Figure 6A**). The patient was refractory to multiple lines of immunosuppression during her management course but ultimately responded to treatment with Tofacitinib, a preferential JAK 1/3 inhibitor, resulting in a down trending of her troponin levels, enabling the weaning off of steroids. Patient 2 had a history of lung cancer and had been started on atezolizumab 3 months prior to presenting with weakness. He was found to have elevated troponins concerning for ICI myocarditis, with **Figure 6B** displaying his troponin trend. He was refractory to steroids and required addition of cellcept, IVIG and ultimately was treated with Tofacitinib, which enabled tapering of his steroids.

### CHIP was also associated with JAK-STAT pathway activation

Four cytokines (FGF2, IL6, IL8 and IL5) emerged as significantly associated with high-risk CHIP score by point biserial correlation (**Supplemental Table 14**). The XGBoost model had an accuracy of 0.809, AUC of 0.803 and F1 score of 0.857. Enriched pathways included JAK-STAT signaling, allograft rejection, type 1 diabetes mellitus, and inflammatory bowel disease (**Supplemental Figure 4**). Our findings regarding enriched pathways associated with both myocarditis and intermediate to high-risk CHIP in our cohort were validated using cytokine data from the Stanford validation cohort (**Supplemental Figure 5).**

### ICI myocarditis is associated with increased activity of pro-inflammatory gene programs in both myeloid and lymphoid lineages in patients with CHIP

Characterization of the activity of gene set signatures across PBMCs in myocarditis and non-myocarditis patients for the MGH, Yale, and Stanford institutional cohorts revealed increased JAK-STAT, cytokine-cytokine interaction signaling and T-cell activation such as in T-cell receptor signaling and Toll-like receptor signaling within cells from myocarditis patients (**Figure 7, Supplemental Figure 6**). Major cell populations mediating this immune response include memory CD4+ T cells, cytotoxic CD8+ NKT cells, and natural killer cells within the lymphoid lineage and non-classical monocytes, plasmacytoid dendritic cells and platelets within the myeloid lineage. When comparing to the pre-steroids sample cohort, our data demonstrated that steroid treatment had only small perturbation effects on cellular signaling patterns in ICI-myocarditis **(Figure 7, Supplemental Figure 7)**. Further, we observed that cells from CHIP patients exhibit increased JAK-STAT signaling, cytokine signaling, and T-cell activation signaling pathways such as T-cell receptor and Toll-like receptor pathways, represented by higher autocorrelation scores in CHIP cell populations **(Supplemental Figure 8)**. In addition to this, CHIP cells exhibited increased cellular signaling of pathways known to be implicated in pro-inflammatory immune response such as NF-kappa B, IL17, MAPK, TNF pathways. Major cell populations mediating this immune response include Memory CD4+, CD8+ NKT cells, and natural killer cells within the lymphoid lineage and classical monocytes, plasmacytoid dendritic cells and platelets within the myeloid lineage **(Supplemental Figure 8)**.

**Figure 7:**
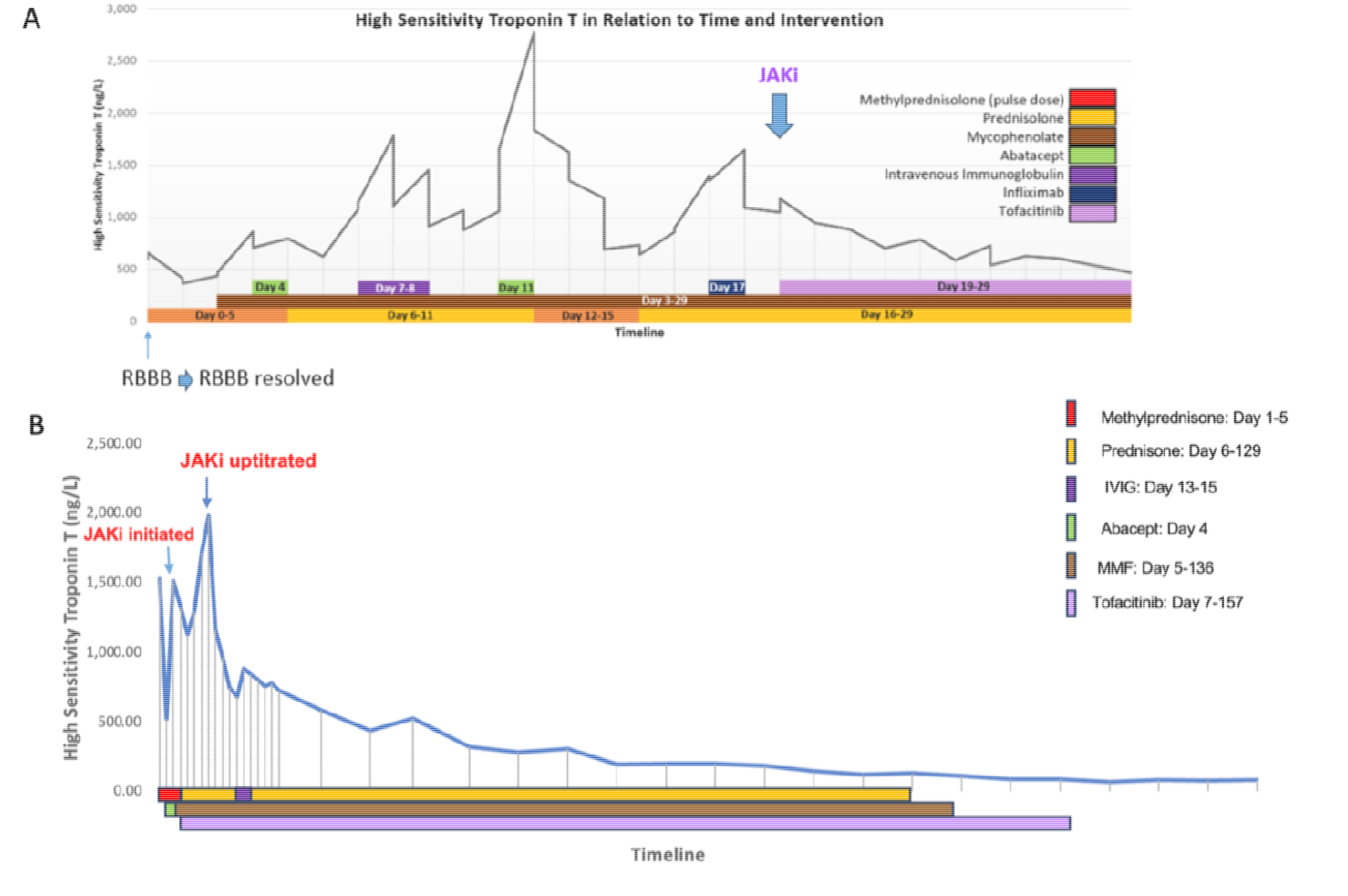
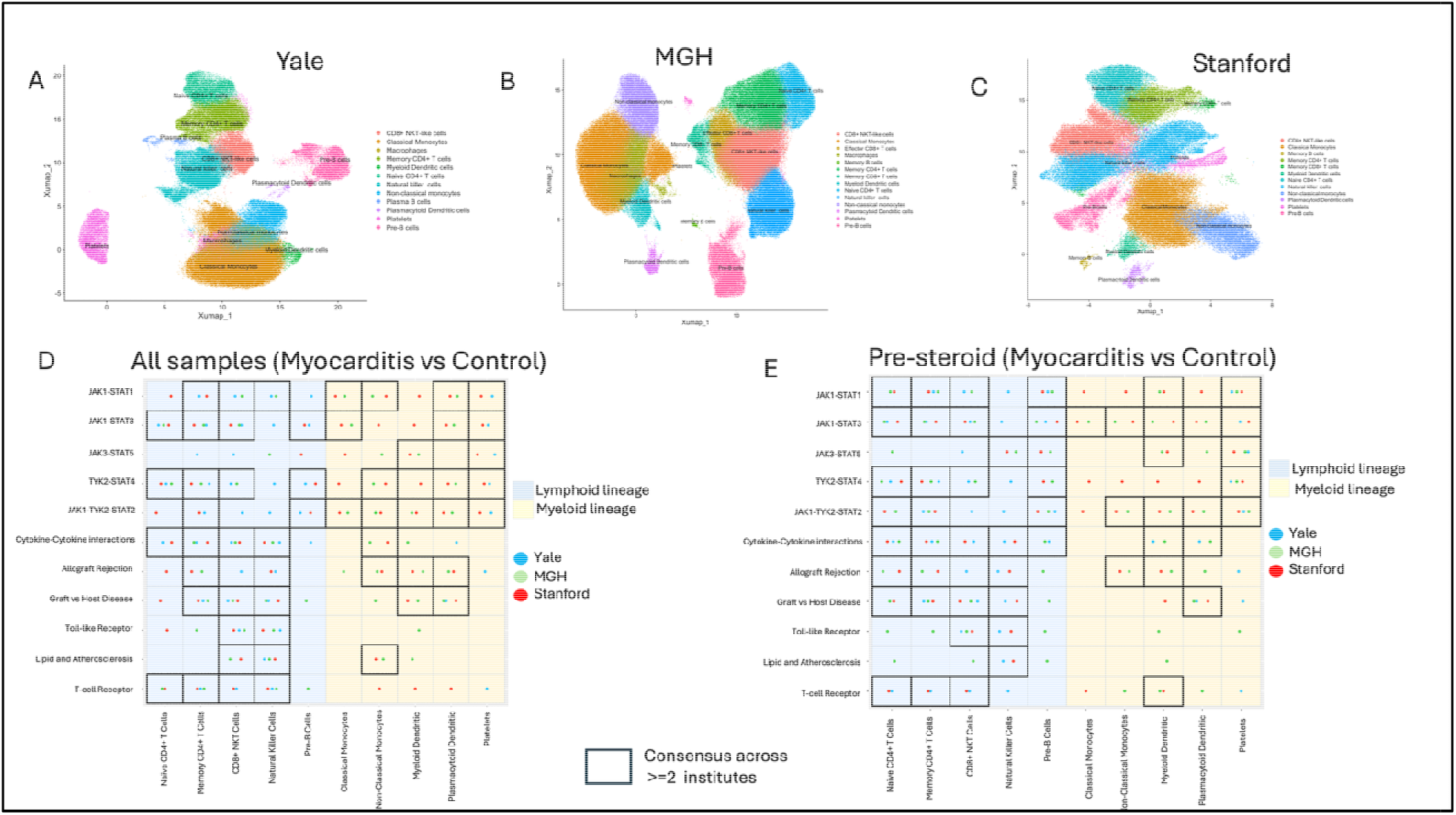
Characterization of activity of gene set signatures in ICI-myocarditis and non-myocarditis patient cohorts using PBMC scRNA-seq data from three institutions. Umap plots show the annotated immune cell clusters for the samples in (A) Yale, (B) MGH, and (C) Stanford. Dot plot (D) shows JAK-STAT signaling and other pro-inflammatory pathways that are upregulated in myocarditis associated immune cells compared to non-myocarditis cells across the lymphoid (blue) and myeloid cell (yellow) lineages when all samples (pre-steroid and post-steroid) were analyzed compared to using only (E) pre-steroid samples.

These findings suggest that CHIP exacerbates the pro-inflammatory immune response in patients with ICI-myocarditis, acting as a secondary inflammatory response that leads to myocardial fibrosis and tissue damage. Similar to our findings in myocarditis and non-myocarditis comparisons, our analysis with CHIP and non-CHIP cell populations demonstrated that steroid treatment only had small perturbation effects on cellular signaling patterns in samples from CHIP-positive patients with ICI-myocarditis **(Figure 8, Supplemental Figure 9)**. Our multi-omics factor analysis (MOFA) of Stanford cohort using both scRNA and CyTOF data identified myocardial chemokines such as CCL5, CCL4, CCL3 and inflammatory cytokines such as TNFSF10, IL1B, IL18, CD40LG as the top variable cytokines differentiating myocarditis and non-myocarditis samples. **Supplemental Results Text 1** provides additional information on the cytokines identified with MOFA analysis. **Supplemental Table 16 and 17** show top features stratified by and myocarditis status and steroid status (pre or all samples (pre+post)). JAK-STAT signaling pathway emerged as one of the top KEGG pathways associated with cytokines exhibiting differences between myocarditis and nonmyocarditis samples (**Supplementary Figure 10**).

**Figure 8:**
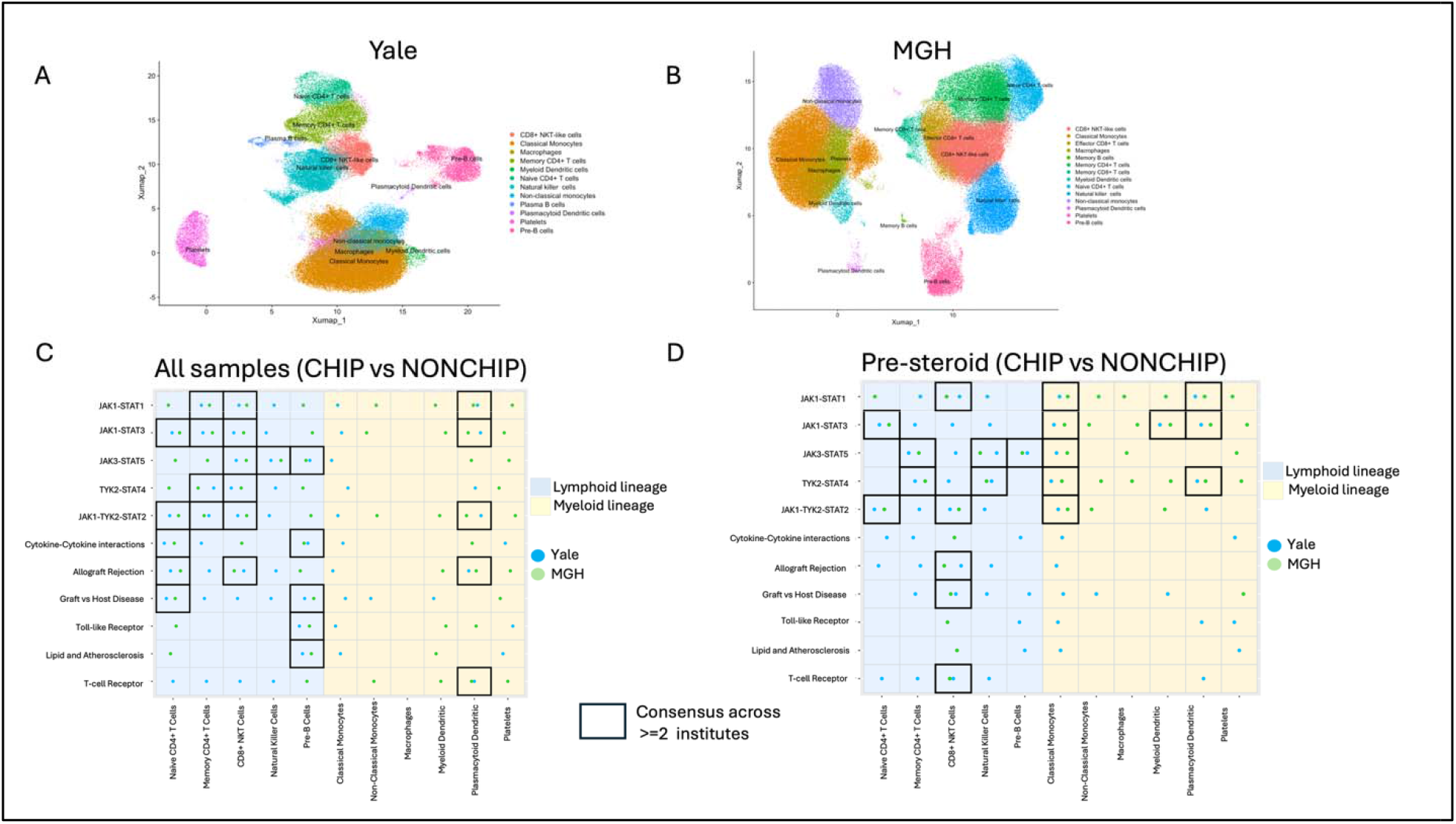
Characterization of activity of gene set signatures in ICI-myocarditis CHIP and ICI-myocarditis NONCHIP patient cohorts using PBMC scRNA-seq data. Umap plots show the annotated immune cell clusters for myocarditis samples in (A) Yale and (B) MGH cohorts. Dot plot (C) shows JAK-STAT signaling and other pro-inflammatory pathways that are upregulated in CHIP associated immune cells compared to NONCHIP cells across the lymphoid (blue) and myeloid cell (yellow) lineages using all samples (pre-steroid and post-steroid) compared to using only (D) pre-steroid samples.

## Discussion

To our knowledge, this is the first study to use cytokine analyses from peripheral blood samples in patients receiving immunotherapy in conjunction with machine learning and explainable artificial intelligence to identify top pathways associated with ICI myocarditis, immunosuppression-resistant ICI myocarditis, and the immune profile of CHIP in ICI myocarditis, with key findings validated in two independent cohorts. As biomarker signatures are needed to promote earlier detection, this sets the stage for development of a biomarker panel to evaluate for ICI myocarditis and immunosuppression resistant ICI myocarditis. Extracting SHAP values from the machine learning model is a unique strength of our manuscript, as it enables us to hypothesize the directionality of the relationship between cytokines and our outcome of interest and allows for hypotheses generation regarding mechanisms through which various cytokines mediate myocarditis.

Several of the identified top cytokine features have known roles in PD-1 and PD-L1 inhibitor mediated toxicity, and in tumor immunology, response to immunotherapy, and cardiovascular disease more generally. Among significant pathways identified, the enrichment of the inflammatory NF-κB pathway and of increased interleukin 17 signaling aligns with prior evidence from *in vitro* models of human cardiomyocytes and mouse models showing a role for increased NF-κB pathway signaling and increased interleukin 17 signaling as drivers for cardiotoxicity from PD-1 blockade respectively (3). The correlation of MIG/CXCL9 with immunosuppression resistant myocarditis and the identification of both MIG/CXCL9 and IP10/CXCL10 as top features in the machine learning model for myocarditis supports existing evidence on the role of expansion of CXCL9/10+ macrophages which recruit CXCR3hi CD8+ effector T cells in mice models of myocarditis and the presence of CXCL9/10+ macrophages in cardiac biopsy staining (18).

Through extracting SHAP values to determine directionality, we can hypothesize the role of a particular cytokine in the context of known literature. MDC/CCL22 (macrophage-derived chemokine) emerged as a highly predictive feature in the immunosuppression resistant myocarditis model and demonstrated a negative correlation with immunosuppression myocarditis. MDC is a known checkpoint and is produced by dendritic cells (DC) facilitating the interaction between DC and regulatory T cells (T_reg_) via the CCR4 receptor (19). Reduced CCL22 expression has been reported to reduce DC and T_reg_ cells leading to an upregulated immune response, as in the case of ICI myocarditis(19).

Similarly, B-cell attracting chemokine (BCA-1/CXCL13) was positively associated with immunosuppression-resistant myocarditis and identified as a top predictive feature in our model.BCA-1 is expressed in B-cell follicles of secondary lymphoid organs and is known for its role as a strong B-cell chemotactic, with emerging evidence supporting its role as a biomarker of immune activation and response to immunotherapy in serous ovarian cancer(20). Its known roles in immune cell recruitment align with its upregulation in the context of immunosuppression resistant myocarditis.

The emergence of JAK-STAT signaling as a significantly enriched pathway in both myocarditis and immunosuppression resistant myocarditis models corroborates the mounting clinical evidence showing efficacy of JAK/STAT inhibitors such as Tofacitinib in treating immunosuppression resistant myocarditis (21). JAK-STAT signaling has also been linked to several other cytokines identified by our model. For instance, some evidence suggests a role for CXCL9 overexpression in promoting JAK1-STAT2 activation (22), a role for JAK-STAT1 signaling in stimulating MDC/CCL22 production by macrophages (23), a role for STAT6 expression in restricting CCL22 production (24), and finally a role for tofacitinib treatment in reducing levels of BCA-1/CXCL13 (25).

Characterization of the JAK-STAT pathways, as well other pro-inflammatory pathways using single-cell RNA-seq in the myocarditis and non-myocarditis cohorts provided mechanistic evidence of the involvement of these pathways in the pathogenesis of ICI-myocarditis. At the single-cell level, module-level JAK-STAT activation reached cross-cohort consensus in CHIP carriers as well as myocarditis comparison, localizing this axis to both lymphoid (memory CD4+, CD8+ NKT) and myeloid (monocytes, dendritic cells, platelets) lineages. Notably, CHIP appeared to amplify pro-inflammatory immune signaling in ICI myocarditis, potentially contributing to the increased risk of myocarditis in cancer patients undergoing immunotherapy.

Future research could assess whether elevation of predictive cytokines identified in our model and enriched in JAK-STAT pathways could be used as biomarkers for predicting need for and potential response to JAK-STAT inhibition to accelerate the right choice of immunosuppressant. Given the emerging role of CHIP as a risk factor for ICI myocarditis, targeting this pathway in patients with CHIP who develop ICI myocarditis may be an effective treatment approach. Although the CHIP risk score has been prognostic for the outcome of death in the UK Biobank cohort (12), its prognostic value in a cancer population remains less clear, and a recent study suggests there is a need for dedicated risk scores in patients with solid cancers (26) as well as in those receiving immunotherapy.

While not statistically significant, the higher rate of TET2 mutations in patients with CHIP who developed ICI myocarditis is noteworthy given the acceleration of atherosclerosis in TET2-deficient mouse models and increased expression of CXCL9 in TET2 deficient monocytes (27). Larger cohort studies are underway to examine significant relationships between CHIP variants and myocarditis.

## Limitations

The major limitation of our study includes the smaller sample size, albeit tempered by having 2 external validation cohorts, and heterogeneity in the cancer population. Our single-cell RNA characterization of the cellular pathways overlap in findings across 3 institutions. However, there are also inconsistencies in the cell cluster patterns associated with activated pathways across the 3 institutions, which we believe could be largely attributed to differences in sample-specific meta-data such as age, co-morbidities, demographics, stage of myocarditis, and technical differences and sample processing techniques. Additional limitations include the observational, associative design, which precludes causal inference; single-timepoint sampling; the finite breadth of the cytokine panels; and the modest number of myocarditis cases, which necessitated synthetic oversampling during model training and warrants cautious interpretation of the reported discrimination metrics.

## Conclusions

Our study used machine learning approaches to identify top cytokine features associated with myocarditis and immunosuppression resistant myocarditis. In line with previous literature, we identified several cytokines predictive of myocarditis and immunosuppression resistant myocarditis, including IP-10/CXCL10, MIG/CXCL9, and BCA-1/CXCL13 among others. JAK-STAT signaling emerged as a significantly enriched pathway across models, adding to the growing body of evidence of its role in myocarditis and the role of JAK-STAT inhibitors in treating immunosuppression resistant myocarditis. Further work is needed to elucidate the mechanisms through which JAK-STAT signaling mediates myocarditis and the role of other identified cytokines in the pathogenesis of the disease. Validation of these cytokine signatures may enable development of clinically actionable biomarker panels to guide risk stratification and immunosuppressive decision-making in cardio-oncology practice.

## Competency in medical knowledge

In patients receiving immunotherapy who develop ICI myocarditis, the JAK-STAT signaling pathway, as well as other pro-inflammatory pathways, including the IL-17 signaling pathway, are significantly upregulated in both myeloid and lymphoid lineages. Clonal hematopoiesis of indeterminate potential further amplifies the JAK-STAT signaling, T-cell activation pathways such as the Toll-like receptor pathways and increases in NF-kappa B and TNF signaling pathways, in accordance with previous research showing that CHIP is a risk factor for ICI myocarditis.

## Translational outlook

Further studies should elucidate the mechanisms through which JAK-STAT signaling mediates myocarditis. Validation studies of predictive cytokine biomarkers would enable early development of a cytokine panel for early detection and risk-stratification of ICI myocarditis development and severity.

## Data Availability

All data produced in the present study are available upon reasonable request to the authors

## Acknowledgements

We thank patients for their participation in the study. N.M and R.J.C contributed equally as first authors. All authors contributed to drafting and editing of the manuscript. We would like to acknowledge Talal El Zarif, Ana Ferrigno Guajardo, and Shaheen Khatua for their contribution to this study.

## Abbreviations

ACE: angiotensin converting enzyme
ARB: angiotensin receptor blocker
CAD: coronary artery disease
CCB: calcium channel blocker
CCUS: clonal cytopenias of undetermined significance
CHIP: clonal hematopoiesis of indeterminate potential
CM: cardiomyopathy
HLD: hyperlipidemia
HTN: hypertension
ICI: immune checkpoint inhibitor
VAF: variant allele fraction
XRT: radiation therapy

## Central Illustrations

serve to succinctly summarize the entire manuscript or at least a major section of the manuscript. Upon acceptance, our in-house medical illustrators will assist in creating the final printable versions of these figures in consultation with the authors and the editors. The purpose of these illustrations is to provide a snapshot of your paper in a single visual, conceptual manner. This illustration must be accompanied by a legend consisting of a title and caption of no more than 50-100 words in total. The title should be no more than 10 words.

## Central illustration

**Title:** Identifying pathways mediating ICI myocarditis through cytokine analyses and multiomic

**Legend:** Our study aimed to identify pathways associated with ICI myocarditis broadly, and with immunosuppression-resistant ICI myocarditis and ICI myocarditis in patients with CHIP, a known ICI myocarditis risk factor, more specifically. To achieve this, we had a two-pronged approach. We performed serum cytokine profiling in patients enrolled in a cardio-oncology cohort across Yale and Stanford and adopted a machine-learning approach (XGBoost and SHAP) to identify cytokines predictive of our outcomes of interest. We simultaneously performed scRNA sequencing on PBMC from three institutional cohorts and further pathway enrichment analyses to identify cell-type contributions of upregulated pathways.

## Table legends

**Table 1:** Demographics and Clinical Characteristics of Overall Cardio oncology Cohort and ICI Myocarditis Subsets

**Table 2:** Cancer Types in Cardio-oncology Cohort Stratified by CHIP status

**Table 3:** Top KEGG pathways and associated cytokines in ICI myocarditis

**Table 4:** Top KEGG pathways and associated cytokines in immunosuppression resistant ICI myocarditis

