## Supplemental Material for "Cytokine Signatures of Immune Checkpoint Inhibitor Myocarditis and the Modifying Role of Clonal Hematopoiesis: A Machine-Learning and Multi-Omics Study"

Supplemental Material Outlines

1. Supplemental Methods Text 1: Patient Demographics, Baseline Comorbidities and Outcomes Data Collection
2. Supplemental Methods Text 2: Assessing CHIP status and Cytokine profiling
3. Supplemental Methods Text 3: Single-cell CHIP inference (CIPHER-CHIP)
4. Supplemental Methods Text 4: XG Boost Machine learning model methodology
5. Supplemental Methods Text 5: Characteristics of patient samples used in multi-omic analyses across the internal Yale and external (MGH and Stanford) cohorts
6. Supplemental Methods Text 6: Yale single-cell RNA-seq library construction and sequencing protocol
7. Supplemental Methods Text 7: single cell RNA-seq data integration and cell clustering Yale, MGH and Stanford cohorts
8. Supplemental Table 1: XGBoost combining with different oversampling techniques
9. Supplemental Table 2: Cohorts evaluated in the XGBoost model, incorporating various oversampling techniques. Cohort /evaluated denotes the total number of profiles entering cross-validation and the number with recorded resistance outcomes in the myocarditis evaluation subset.
10. Supplemental Table 3: Summary of the 71 cytokine tests per endpoint
11. Supplemental Table 4: Table showing the myocarditis cytokine associations using the 272 original profiles with 52 outcome-positive. Two-sided point-biserial correlation analysis was performed. Bold entries show BH q < 0.05. Profile-level P values are not cluster-adjusted; group-mean sensitivity retains the same selected assays.
12. Supplemental Table 5: Table showing the Resistance cytokine associations among myocarditis cases using the 50 original profiles with 24 outcome-positive. Two-sided point-biserial correlation analysis was performed. None of the 71 resistance associations retains BH q < 0.05.
13. Supplemental Table 6: Table listing the set of genes used for characterizing cellular pathways in ICI-myocarditis using scRNA-seq data
14. Supplemental Methods Text 8: Pathway Enrichment Characterization methodology
15. Supplemental Methods Text 9: MOFA data processing methodology
16. Supplemental Table 7: Table showing number of CHIP mutations among patients on ICI stratified by myocarditis outcome
17. Supplemental Figure 1: Bar graph showing distribution of entire cohort by number of CHIP mutations
18. Supplemental Table 8: Table of CHIP Variant Frequencies a) Among all CHIP patients b) Among CHIP Patients on ICI with myocarditis c) Among CHIP Patients on ICI who did not develop myocarditis.
19. Supplemental Table 9: Cancer Subtypes Among ICI recipients stratified by CHIP Status
20. Supplemental Table 10: Demographics and Clinical Characteristics of 12 MSKCC Patients and 6 Non-Cancer Controls
21. Supplemental Figure 2: Correlation plots of individual cytokines with myocarditis
22. Supplemental Figure 3: Correlation plots of individual cytokines with immunosuppression resistant myocarditis
23. Supplemental Table 11: Cytokines identified by XGBoost and Corresponding Gene Names
24. Supplemental Table 12: All significant pathways for myocarditis model
25. Supplemental Table 13: All significant pathways for immunosuppression resistant myocarditis model
26. Supplemental Table 14: Cytokines significantly associated with high risk CHIP score by point biserial correlation
27. Supplemental Figure 4: SHAP model [RJ1] (left) and graph of top implicated KEGG pathways (right) for top cytokine features associated with high and intermediate risk CHIP
28. Supplemental Figure 5: Graph of top 10 implicated KEGG pathways associated with both ICI myocarditis (A) and intermediate to high risk CHIP (B) using cytokine features from the Stanford validation cohort
29. Supplemental Figure 6: A, B,C Umap of annotated cell populations in the MGH, YALE, and Stanford cohorts. C, D, E Heatmap illustrating differences in the autocorrelation scores between Myocarditis and Non-myocarditis (control) cells, demonstrating the cell activity of gene programs in these two conditions for all PBMC scRNA-seq samples (pre-steroid and post-steroid samples) from MGH, YALE, and Stanford cohorts.
30. Supplemental Table 15: Demographics and Clinical Characteristics of 19 patients with PBMC tissue samples comprising internal Yale cohort in multiomics analysis
31. Supplemental Figure 7: A, B,C Umap of annotated cell populations in the MGH, YALE, and Stanford cohorts. C, D, E Heatmap illustrating differences in the autocorrelation scores between Myocarditis and Non-myocarditis (control) cells, demonstrating the cell activity of gene programs in these two conditions for pre-steroid PBMC scRNA-seq samples from MGH, YALE, and Stanford cohorts.
32. Supplemental Figure 8: A, B Umap of annotated cell populations in the MGH and YALE cohort. C, D Heatmap illustrating the differences in autocorrelation scores between Myocarditis CHIP and Myocarditis NONCHIP cells, demonstrating the cell activity of gene programs in these two conditions for all PBMC scRNA-seq samples (pre-steroid and post-steroid samples) from MGH and YALE cohorts.
33. Supplemental Figure 9: Umap of annotated cell populations in the MGH and YALE cohort. C, D Heatmap illustrating the differences in autocorrelation scores between Myocarditis CHIP and Myocarditis NONCHIP cells, demonstrating the cell activity of gene programs in these two conditions for pre-steroid PBMC scRNA-seq samples from MGH and YALE cohorts.
34. Supplemental Figure 10: Cytoscape visualization of the graph structure of cytokine-cytokine interaction signaling showing the high connectivity cytokines (represented by high degree nodes (dark colored central nodes)).
35. Supplemental Results Text 1: MOFA results
36. Supplemental Table 16. Top features in the Stanford cohort, myocarditis-stratified.
37. Supplemental Table 17. Pre-steroid versus pre+post-steroid top features for the Stanford cohort, stratified by myocarditis status. Stanford is the cohort with matched CyTOF and scRNA-seq data.
38. Supplemental Figure 11: Top KEGG pathways associated with the cytokines that differed between myocarditis and nonmyocarditis samples in the Stanford cohort (top features listed in Supplemental Table 16).

**Supplemental Methods Text 1:** **Patient Demographics, Baseline Comorbidities and Outcomes Data Collection**

Structured chart review of outpatient and inpatient records was performed to determine patient demographics, oncologic history, baseline comorbidities and cardiac function and outcomes. Collected variables were cancer and cancer treatment history, baseline comorbidities, cardiac medications, and baseline cardiac imaging, defined as echocardiogram (echo) or cardiac MRI (cMRI) within one month of blood draw. Outcomes of interest included the development of myocarditis, immunosuppression resistant myocarditis, new cardiomyopathy, diastolic dysfunction, and death. Myocarditis was defined based on follow up cMRI imaging as per the International Cardio-Oncology (IC-OS) definition for ICI myocarditis, based on the major criteria (diagnostic cMRI per the modified Lake Louise criteria) or the minor criteria combining clinical and cMRI findings (1). Immunosuppression resistant myocarditis was defined as patients requiring adjunct immunosuppression agents on top of the standard pulse dose steroids followed by steroid taper. New cardiomyopathy was defined as EF <50% or drop of 10% from baseline for patients with prior cardiomyopathy. For patients with myocarditis, we recorded the timing of blood draw with respect to the start of immunosuppression, and classified patients accordingly to pre- and post-steroids. Only pre-steroid samples were used in the machine learning models for cytokine analysis to evaluate cytokine signatures associated with ICI myocarditis and immunosuppression resistant ICI myocarditis. For multi-center cytokine profiling using MOFA to compare pre and post steroids, myocarditis vs non myocarditis, CHIP vs no CHIP, all samples were used. Diastolic dysfunction (grades 1-3) was defined per echocardiographic findings on follow up imaging (2).

**Supplemental Methods Text 2: Assessing CHIP status and Cytokine profiling**

CHIP variants were annotated across established CHIP genes, including DNMT3A, TET2, and IDH2, and variant burden per individual was recorded. Additionally, protein levels of 71 serum cytokines were evaluated via a commercially available multiplex immunoassay (Eve Technologies); this serum panel is distinct from the 55-gene targeted panel used for the single-cell analyses.

**Supplemental Methods Text 3: Single-cell CHIP inference (CIPHER-CHIP)**

CIPHER-CHIP was not used as a stand-alone DNA variant caller, since scRNA-seq provides sparse, transcript-biased coverage of somatic mutations,. It integrated single-cell SNV detection, expressed-allele quantification, bulk DNA-derived CHIP calls, and donor-level clonal architecture to estimate, for each cell, the probability of belonging to a CHIP-mutant clone. Aligned scRNA-seq reads (Cell Ranger (3) underwent standard quality control, ambient-RNA and doublet removal, normalization, and batch-corrected integration. Single-cell SNVs were genotyped with Monopogen (4) using external haplotype reference panels and known CHIP alleles defined from matched bulk and whole-exome sequencing were quantified across cell barcodes using cellSNP-lite (5) and VarTrix (GitHub–10XGenomics/vartrix: Single-Cell Genotyping Tool), with independent confirmation by GATK ASEReadCounter, bcftools, and samtools (6,7). Candidate variants were restricted to canonical CHIP driver genes (including DNMT3A, TET2, ASXL1, JAK2, PPM1D, TP53, SF3B1, SRSF2, and IDH1/2) and annotated against population and somatic databases (8). Directly mutation-positive cells served as high-confidence seed cells; because many clone members lack mutant read coverage owing to transcript dropout, latent clone membership was inferred across the immune atlas using a donor-aware probabilistic graph model (implemented in Pyro/PyMC (9,10)) that combined direct allele evidence, dropout probability, mutated-gene expression, transcriptional similarity to seed cells, and donor-level bulk variant allele fraction, while adjusting for donor, cell-type, batch, age, sex, and sequencing-depth covariates. Cells were classified as CHIP-positive, CHIP-negative, or uncertain using calibrated posterior thresholds, and clone assignments were assessed for concordance with bulk DNA variant allele fraction and across independent allele callers.

**Supplemental Methods Text 4:** XG Boost Machine learning model methodology

XG Boost , a highly successful ensemble method for tabular data, even outperforming deep neural networks (11). Taking into consideration the small cohort and the imbalance in our dataset (where only about 19% of instances have positive labels), we applied several previously validated optimization techniques to improve the model's performance (12,13). These techniques include hyperparameter tuning, feature normalization, and implementation of oversampling techniques for the minority classes. We utilized the Synthetic Minority Oversampling Technique (SMOTE) and the Adaptive Synthetic (ADASYN) specifically(12,13). These techniques select a random sample from the minority class, identify its k nearest neighbors, and form new synthetic samples by interpolation between the chosen sample and its neighbors. The process helps to overcome the class imbalance problem by increasing the size of the minority class. To explore feature interpretability, we extracted the SHAP values after training XGBoost which provide information on the direction of the relationship between cytokine and outcome (14). For each task, we employed 5-fold cross-validation approach and report the prediction area-under-the-curve (AUC), accuracy, and F1-score. Instead of random splitting, we opted for stratified 5-fold cross-validation to ensure that the ratio between the target classes remains consistent in each fold as it is in the full dataset. We built XGBoost models using 219 cytokine profiles as input data to identify cytokine signatures associated with myocarditis and immunosuppression resistant myocarditis. The correlation plots (**Supplemental Figure 2 and 3)** were generated from two endpoint-specific datasets constructed from the same cohort of 77 cytokine profiles, comprising 66 profiles coded as ICI-treated and 11 coded as non-ICI. The same 71 cytokine measurements were extracted for both datasets. All 77 profiles had recorded labels for both outcomes. The myocarditis dataset contained 30 positive and 47 negative profiles. The immunosuppression-resistance dataset contained 9 positive and 68 negative profiles; the latter included 47 non-myocarditis profiles. Cytokine–outcome associations were evaluated using point-biserial correlation, equivalent to Pearson correlation between a continuous measurement and a binary outcome. Correlation coefficients and two-sided, unadjusted P values were calculated using scipy.stats.pointbiserialr.

#### **Metric definitions**

AUC (Area under the curve) summarizes ranking across sensitivity and false-positive rate. AUPRC (Area Under the Precision-Recall Curve) is the trapezoidal area under the precision–recall curve. AP uses the stepwise precision-weighted recall increments and is reported separately because it differs from trapezoidal AUPRC, particularly for tied or constant predictions. Accuracy = (TP + TN)/N and positive-class F1 = 2TP/(2TP + FP + FN); zero-denominator F1 was assigned zero.

### **Myocarditis XGBoost comparisons**

Fixed XGBoost used prespecified hyperparameters, including 100 trees, a maximum tree depth of 6, and a learning rate of 0.3. Optimized XGBoost selected among 13 predefined hyperparameter configurations using three-fold inner cross-validation, with additional class-weighting options for models without augmentation. Optimized XGBoost + SMOTE and Optimized XGBoost + ADASYN incorporated their respective synthetic minority oversampling methods and additionally evaluated minority-to-majority sampling ratios of 0.5 and 1.0. Synthetic observations were generated exclusively within training partitions. Classification thresholds were selected using inner out-of-fold predictions to maximize positive-class F1. Performance was evaluated using five-fold outer cross-validation repeated three times, with held-out observations excluded from model and threshold selection. Please note that SMOTE/ADASYN oversampling was performed after the split, and used training data only, to avoid data leakage. (**Supplemental Table 1**)

**Supplemental Table 1:** XGBoost combining with different oversampling techniques

| **Procedure** | **AUC** | **AUPRC** | **Accuracy** | **F1** |
| --- | --- | --- | --- | --- |
| Optimized XGBoost | 0.915 | 0.705 | 0.866 | 0.692 |
| Optimized XGBoost + SMOTE | 0.918 | 0.715 | 0.869 | 0.713 |
| Optimized XGBoost + ADASYN | 0.916 | 0.697 | 0.866 | 0.705 |

### **Immunosuppression resistance**

The cohorts of 52, 90 and 272 profiles respectively assigned 2, 33 and 196 missing resistance labels to zero in training. These are unverified sensitivity assumptions; all evaluated outcomes were recorded. Each cohort retained the same 24 recorded resistant profiles. The 90-profile cohort required independently recorded ICI therapy (**Supplemental Table 2**).

**Supplemental Table 2:** Cohorts evaluated in the XGBoost model, incorporating various oversampling techniques. Cohort /evaluated denotes the total number of profiles entering cross-validation and the number with recorded resistance outcomes in the myocarditis evaluation subset.

| **Model** | **Cohort / evaluated** | **AUC** | **AUPRC** | **Accuracy** | **F1** |
| --- | --- | --- | --- | --- | --- |
| Optimized XGBoost | 52 / 50 | 0.702 | 0.707 | 0.660 | 0.706 |
| XGBoost + SMOTE | 90 / 48 | 0.773 | 0.787 | 0.694 | 0.732 |
| XGBoost + ADASYN | 272 / 50 | 0.798 | 0.797 | 0.680 | 0.717 |

### **Exploratory cytokine associations**

#### **Association methods**

Each of 71 unlogged cytokines was tested against the binary recorded endpoint using a two-sided point-biserial correlation, equivalent to Pearson correlation with a binary outcome. Positive r indicates higher values in outcome-positive profiles. Analyses used original observed rows in the frozen cohorts; synthetic observations, oversampled observations, fitted probabilities and repeated cross-validation rows were excluded. No new log transformation, scaling or imputation was applied. All 71 assays had finite, variable values. These are univariate associations without clinical covariate adjustment or model-specific feature-importance interpretation.

Benjamini–Hochberg (BH) q values were calculated across all 71 cytokines separately for each endpoint (**Supplemental Table 3**). Complete results are reported in **Supplemental Tables 4** **and** **5**, ordered by unadjusted P. Nominal P < 0.05 alone was considered exploratory; bold entries indicate q < 0.05. Model variants evaluated on the same cohort share this association table.

**Supplemental Table 3:** Summary of the 71 cytokine tests per endpoint

| **Endpoint** | **Profiles** | **P < 0.05** | **BH q < 0.05** |
| --- | --- | --- | --- |
| Myocarditis | 272 | 21 | 11 |
| Resistance among myocarditis cases | 50 | 8 | 0 |

For myocarditis, 11 cytokines retained q < 0.05: MIG/CXCL9, BCA-1, MIP-1a, IL-1RA, IL-27, IL-15, IL-1B, IP-10, IL-2, IL-18 and MDC. MIG/CXCL9 had the smallest P value (r = 0.367; P = 4.15 × 10⁻¹⁰; q = 2.95 × 10⁻⁸). For resistance, eight cytokines had nominal P < 0.05, but none retained q < 0.05; the smallest P value was for MIG/CXCL9 (r = 0.406; P = 0.00345; q = 0.245).

The 272 myocarditis profiles represent 269 recorded groups. Conventional profile-level P values assume independence and are not cluster-adjusted. Three groups each had two profiles with different cytokine values and consistent negative labels. A sensitivity analysis averaging cytokines within each group produced 269 group means, including 52 positive groups, and retained exactly the same 21 nominal and 11 BH-selected cytokines. Post-immunosuppression profiles remained in both cohorts; these associations do not establish pretreatment biomarkers, causality or independent prediction performance.

**Supplemental Table 4:** Table showing the myocarditis cytokine associations using the 272 original profiles with 52 outcome-positive. Two-sided point-biserial correlation analysis was performed. Bold entries show BH q < 0.05. Profile-level P values are not cluster-adjusted; group-mean sensitivity retains the same selected assays.

| **Cytokine** | **Correlation r** | **Two-sided P** | **BH q** |
| --- | --- | --- | --- |
| **MIG/CXCL9** | **0.367227** | **4.151e-10** | **2.947e-08** |
| **BCA-1** | **0.275319** | **4.043e-06** | **1.435e-04** |
| **MIP-1a** | **0.247764** | **3.596e-05** | **8.511e-04** |
| **IL-1RA** | **0.214234** | **3.732e-04** | **6.624e-03** |
| **IL-27** | **0.203442** | **7.377e-04** | **1.048e-02** |
| **IL-15** | **0.192737** | **1.403e-03** | **1.660e-02** |
| **IL-1B** | **0.189361** | **1.706e-03** | **1.731e-02** |
| **IP-10** | **0.185714** | **2.101e-03** | **1.865e-02** |
| **IL-2** | **0.181339** | **2.683e-03** | **2.117e-02** |
| **IL-18** | **0.166209** | **6.002e-03** | **4.261e-02** |
| **MDC** | **-0.163163** | **7.004e-03** | **4.521e-02** |
| IFNg | 0.150709 | 1.283e-02 | 7.594e-02 |
| FGF-2 | 0.147610 | 1.483e-02 | 8.068e-02 |
| IL-6 | 0.145431 | 1.638e-02 | 8.068e-02 |
| IL-17E/IL-25 | 0.143142 | 1.817e-02 | 8.068e-02 |
| IL-12p70 | 0.143132 | 1.818e-02 | 8.068e-02 |
| GM-CSF | 0.132678 | 2.869e-02 | 1.198e-01 |
| PDGF-AB/BB | 0.128515 | 3.413e-02 | 1.309e-01 |
| IL-17A | 0.127889 | 3.502e-02 | 1.309e-01 |
| IL-7 | 0.123464 | 4.189e-02 | 1.427e-01 |
| IL-1a | 0.123279 | 4.220e-02 | 1.427e-01 |
| IL-13 | 0.116499 | 5.498e-02 | 1.634e-01 |
| TGFa | 0.116321 | 5.535e-02 | 1.634e-01 |
| Fractalkine | 0.116195 | 5.562e-02 | 1.634e-01 |
| IL-9 | 0.115299 | 5.755e-02 | 1.634e-01 |
| TNFa | 0.112850 | 6.309e-02 | 1.723e-01 |
| PDGF-AA | 0.109256 | 7.202e-02 | 1.882e-01 |
| IL-10 | 0.107698 | 7.620e-02 | 1.882e-01 |
| IL-5 | 0.107457 | 7.686e-02 | 1.882e-01 |
| M-CSF | 0.105030 | 8.381e-02 | 1.976e-01 |
| IL-4 | 0.104212 | 8.626e-02 | 1.976e-01 |
| TNFb | 0.098582 | 1.047e-01 | 2.324e-01 |
| Eotaxin-2 | -0.090780 | 1.353e-01 | 2.912e-01 |
| TSLP | 0.088985 | 1.433e-01 | 2.992e-01 |
| TRAIL | -0.084846 | 1.629e-01 | 3.299e-01 |
| VEGF-A | 0.083973 | 1.673e-01 | 3.299e-01 |
| MCP-1 | -0.078099 | 1.991e-01 | 3.782e-01 |
| MIP-1b | 0.076963 | 2.057e-01 | 3.782e-01 |
| MCP-3 | 0.076625 | 2.078e-01 | 3.782e-01 |
| MCP-2 | -0.071806 | 2.379e-01 | 4.159e-01 |
| IL-8 | 0.071459 | 2.402e-01 | 4.159e-01 |
| CTACK | 0.070306 | 2.478e-01 | 4.190e-01 |
| SCF | 0.067241 | 2.691e-01 | 4.443e-01 |
| IFN-a2 | 0.062231 | 3.065e-01 | 4.946e-01 |
| IL-33 | 0.060822 | 3.176e-01 | 5.011e-01 |
| Eotaxin | -0.058875 | 3.334e-01 | 5.145e-01 |
| MCP-4 | -0.055609 | 3.609e-01 | 5.452e-01 |
| EGF | -0.052473 | 3.887e-01 | 5.749e-01 |
| ENA-78 | -0.046760 | 4.425e-01 | 6.411e-01 |
| IL-28A | 0.041414 | 4.964e-01 | 7.026e-01 |
| IL-16 | 0.040618 | 5.047e-01 | 7.026e-01 |
| IL-21 | 0.038625 | 5.259e-01 | 7.180e-01 |
| TPO | 0.030883 | 6.121e-01 | 8.200e-01 |
| SDF-1a+b | 0.029851 | 6.240e-01 | 8.205e-01 |
| IL-20 | -0.027445 | 6.523e-01 | 8.420e-01 |
| LIF | -0.023563 | 6.988e-01 | 8.717e-01 |
| 6CKine | 0.023372 | 7.012e-01 | 8.717e-01 |
| IL-3 | -0.021906 | 7.191e-01 | 8.717e-01 |
| IL-23 | -0.020930 | 7.311e-01 | 8.717e-01 |
| sCD40L | 0.020037 | 7.422e-01 | 8.717e-01 |
| Eotaxin-3 | -0.019494 | 7.489e-01 | 8.717e-01 |
| MIP-1d | 0.013820 | 8.205e-01 | 9.250e-01 |
| G-CSF | 0.013797 | 8.208e-01 | 9.250e-01 |
| I-309 | -0.012283 | 8.402e-01 | 9.321e-01 |
| TARC | 0.011204 | 8.541e-01 | 9.329e-01 |
| IL-17F | -0.007024 | 9.082e-01 | 9.625e-01 |
| IL-22 | 0.007017 | 9.083e-01 | 9.625e-01 |
| RANTES | -0.005208 | 9.319e-01 | 9.717e-01 |
| FLT-3L | 0.003853 | 9.496e-01 | 9.717e-01 |
| GROa | -0.003206 | 9.580e-01 | 9.717e-01 |
| IL-12p40 | -0.001217 | 9.841e-01 | 9.841e-01 |

**Supplemental Table 5:** Table showing the Resistance cytokine associations among myocarditis cases using the 50 original profiles with 24 outcome-positive. Two-sided point-biserial correlation analysis was performed. None of the 71 resistance associations retains BH q < 0.05.

| **Cytokine** | **Correlation r** | **Two-sided P** | **BH q** |
| --- | --- | --- | --- |
| MIG/CXCL9 | 0.405834 | 3.455e-03 | 2.453e-01 |
| FLT-3L | -0.363018 | 9.568e-03 | 2.972e-01 |
| MIP-1a | -0.350580 | 1.256e-02 | 2.972e-01 |
| MDC | -0.323359 | 2.199e-02 | 3.387e-01 |
| IL-6 | -0.317167 | 2.481e-02 | 3.387e-01 |
| MIP-1b | 0.309696 | 2.862e-02 | 3.387e-01 |
| GROa | -0.280849 | 4.820e-02 | 4.010e-01 |
| LIF | -0.279271 | 4.952e-02 | 4.010e-01 |
| MCP-1 | -0.272672 | 5.539e-02 | 4.010e-01 |
| IL-17F | -0.271513 | 5.648e-02 | 4.010e-01 |
| VEGF-A | -0.246003 | 8.505e-02 | 4.252e-01 |
| MCP-4 | -0.235234 | 1.001e-01 | 4.252e-01 |
| BCA-1 | 0.235100 | 1.003e-01 | 4.252e-01 |
| IL-8 | -0.231252 | 1.061e-01 | 4.252e-01 |
| GM-CSF | -0.226241 | 1.141e-01 | 4.252e-01 |
| Eotaxin | -0.222701 | 1.201e-01 | 4.252e-01 |
| I-309 | -0.220998 | 1.230e-01 | 4.252e-01 |
| IL-12p40 | -0.217446 | 1.293e-01 | 4.252e-01 |
| IL-4 | -0.211395 | 1.406e-01 | 4.252e-01 |
| IL-17E/IL-25 | -0.207529 | 1.481e-01 | 4.252e-01 |
| MCP-2 | -0.207055 | 1.491e-01 | 4.252e-01 |
| IL-9 | -0.205159 | 1.529e-01 | 4.252e-01 |
| IL-16 | -0.204463 | 1.544e-01 | 4.252e-01 |
| IL-7 | -0.202674 | 1.581e-01 | 4.252e-01 |
| IL-5 | -0.195874 | 1.728e-01 | 4.252e-01 |
| EGF | -0.191387 | 1.830e-01 | 4.252e-01 |
| IFNg | -0.189668 | 1.871e-01 | 4.252e-01 |
| PDGF-AA | -0.189343 | 1.879e-01 | 4.252e-01 |
| TNFb | -0.187597 | 1.920e-01 | 4.252e-01 |
| Fractalkine | -0.186251 | 1.953e-01 | 4.252e-01 |
| IL-2 | -0.180579 | 2.095e-01 | 4.252e-01 |
| IL-1a | -0.180186 | 2.105e-01 | 4.252e-01 |
| MCP-3 | -0.179748 | 2.116e-01 | 4.252e-01 |
| IL-13 | -0.179493 | 2.123e-01 | 4.252e-01 |
| IL-1RA | -0.179098 | 2.133e-01 | 4.252e-01 |
| IL-12p70 | -0.177258 | 2.181e-01 | 4.252e-01 |
| TNFa | -0.174605 | 2.252e-01 | 4.252e-01 |
| TRAIL | -0.173744 | 2.276e-01 | 4.252e-01 |
| FGF-2 | -0.159407 | 2.688e-01 | 4.714e-01 |
| IL-22 | -0.159177 | 2.695e-01 | 4.714e-01 |
| IL-1B | -0.158302 | 2.722e-01 | 4.714e-01 |
| IL-33 | -0.148956 | 3.019e-01 | 4.819e-01 |
| IL-3 | -0.148677 | 3.028e-01 | 4.819e-01 |
| M-CSF | -0.148666 | 3.028e-01 | 4.819e-01 |
| TGFa | -0.145728 | 3.126e-01 | 4.819e-01 |
| TPO | -0.144089 | 3.181e-01 | 4.819e-01 |
| IL-23 | -0.143836 | 3.190e-01 | 4.819e-01 |
| MIP-1d | 0.139321 | 3.346e-01 | 4.870e-01 |
| PDGF-AB/BB | -0.138385 | 3.379e-01 | 4.870e-01 |
| TSLP | -0.136941 | 3.430e-01 | 4.870e-01 |
| IL-18 | -0.130447 | 3.666e-01 | 5.103e-01 |
| IL-27 | -0.126770 | 3.803e-01 | 5.193e-01 |
| IL-17A | -0.121738 | 3.997e-01 | 5.313e-01 |
| IFN-a2 | -0.120586 | 4.042e-01 | 5.313e-01 |
| Eotaxin-2 | 0.118723 | 4.115e-01 | 5.313e-01 |
| IL-15 | -0.104090 | 4.719e-01 | 5.983e-01 |
| IL-28A | -0.097761 | 4.994e-01 | 6.211e-01 |
| IL-20 | -0.095954 | 5.074e-01 | 6.211e-01 |
| ENA-78 | 0.093277 | 5.194e-01 | 6.250e-01 |
| G-CSF | -0.084034 | 5.618e-01 | 6.648e-01 |
| IP-10 | -0.073392 | 6.125e-01 | 7.006e-01 |
| IL-10 | -0.071985 | 6.193e-01 | 7.006e-01 |
| 6CKine | 0.065140 | 6.531e-01 | 7.006e-01 |
| Eotaxin-3 | 0.064591 | 6.559e-01 | 7.006e-01 |
| IL-21 | 0.063640 | 6.606e-01 | 7.006e-01 |
| TARC | -0.062761 | 6.650e-01 | 7.006e-01 |
| CTACK | -0.062616 | 6.658e-01 | 7.006e-01 |
| sCD40L | -0.061565 | 6.710e-01 | 7.006e-01 |
| SDF-1a+b | 0.054390 | 7.076e-01 | 7.281e-01 |
| RANTES | -0.032634 | 8.220e-01 | 8.337e-01 |
| SCF | 0.025278 | 8.617e-01 | 8.617e-01 |

.

**Supplemental Methods Text 5**: Characteristics of patient samples used in multi-omics analyses across the internal Yale and external (MGH and Stanford) cohorts

Our internal Yale post-ICI myocarditis cohort comprises 8 CHIP-positive patients with TET2 (n=2), DNMT3A (n=4), and composite (NOTCH2, DNMT3A, TET2, JAK2) (n=2) mutations, and 5 CHIP-negative patients. Among the 13 PBMC samples in the Yale cohort, 11 samples were collected prior to steroid exposure (7 CHIP-positive, 4 CHIP-negative), while 2 are post-steroid samples (1 CHIP-positive, 1 CHIP-negative). Our Yale post-ICI non-myocarditis cohort consists of three CHIP-positive patients with PPM1D (n=1) and DNMT3A (n=2) mutations, and three CHIP-negative patients (**Supplemental Table** **15**) .

The Stanford post-ICI cohort (15) comprises of 8 post-ICI myocarditis (6 pre-steroid samples). The Stanford post-ICI non-myocarditis cohort consists of 17 samples (15 pre-steroid samples).

The MGH post-ICI myocarditis cohort (16) comprises of 12 CHIP-positive patients, and 25 CHIP-negative patients. Samples designated as post and pre-steroid samples in the published meta-data were included in the post-ICI myocarditis cohort. Among the 37 myocarditis samples, 17 are pre-steroid samples (5 CHIP-positive, 12 CHIP-negative). The MGH non-myocarditis cohort (samples designated as control) consists of 9 CHIP-positive, and 17 CHIP-negative patients. Pre-ICI samples were filtered out from the MGH cohort before downstream analysis.

**Supplemental Methods Text 6:** Yale single-cell RNA-seq library construction and sequencing protocol

We followed a standard single-cell protocol for construction and sequencing of 10X Genomic Single Cell/Nuclei GEM-X 3’ RNA-Seq libraries. Raw sequencing reads were pre-processed using 10x Genomics Cell Ranger 9.0.1 and FASTQ reads were aligned to the human reference genome (GRCh38) and count unique molecular identifiers (UMIs) to generate cell*gene count matrix. We followed the Yale single-cell RNA-seq library construction and sequencing protocol as below:

**Sample Preparation**. The first step of scRNA-Seq involves preparation of the single cell suspension, which would differ depending on the starting sample types. With samples that would perform CITE-Seq (Cellular Indexing of Transcriptomes and Epitopes by Sequencing) together with transcriptomics analysis, the cells are then bound to the oligonucleotide barcoded antibodies of surface proteins. Cell surface proteins can be labeled using a specific protein binding molecule, such as an antibody conjugated to a Feature Barcode oligonucleotide. Additional nuclei isolation would be needed for single nuclei transcriptomics analysis.

**GEM Generation and Barcoding.** Single cell/nuclei suspension in RT Master Mix is loaded on the Single Cell A Chip and partition with a pool of about 3,600,000 barcoded gel beads to form nanoliter-scale Gel Beads-In-Emulsions (GEMs). With each gel bead, it has primers containing (i) an Illumina® R1 sequence (read 1 sequencing primer), (ii) a 16 nt 10x Barcode, (iii) a 12 nt Unique Molecular Identifier (UMI), and (iv) a poly-dT primer sequence (30nt) . Upon dissolution of the Gel Beads in a GEM, the primers are released and mixed with cell lysate and Master Mix. Incubation of the GEMs then produces barcoded, full-length cDNA from poly-adenylated mRNA.

**Post GEM-RT Cleanup, cDNA Amplification and library construction**. Silane magnetic beads are used to remove leftover biochemical reagents and primers from the post GEM reaction mixture. Full-length, barcoded cDNA is then amplified by PCR to generate sufficient mass for library construction. Enzymatic Fragmentation and Size Selection are used to optimize the cDNA amplicon size prior to library construction. R1 (read 1 primer sequence) are added to the molecules during GEM incubation. P5, P7, a sample index, and R2 (read 2 primer sequence) are added during library construction via End Repair, A-tailing, Adaptor Ligation, and PCR. The final libraries contain the P5 and P7 primers used in Illumina bridge amplification.

**Sequencing libraries.** The Single Cell GEM-X 3’ Protocol produces Illumina-ready sequencing libraries. A Single Cell 3’ Library comprises standard Illumina paired-end constructs which begin and end with P5 and P7. The Single Cell 3’ 16 bp 10x Barcode and 12 bp UMI are encoded in Read 1, while Read 2 is used to sequence the cDNA fragment (91bp). Sequencing a Single Cell 3’ Library produces a standard Illumina BCL data output folder. The BCL data will include the paired-end Read 1 (containing the 16 bp 10x™ Barcode and 12 bp UMI) and Read 2 and the sample index in the i7 index read. Minimum sequencing depth is 20,000 read pairs per cell.

**Single Cell 3’ library analysis.** The Cell Ranger™ analysis pipelines perform secondary analysis and visualization. In addition to performing standard analysis steps such as demultiplexing, alignment, and gene counting, Cell Ranger™ leverages the 10x Barcodes to generate expression data with single-cell resolution. This data type enables applications including cell clustering, cell type classification, and differential gene expression at a scale of hundreds to millions of cells.

**Supplemental Methods Text 7:** single cell RNA-seq data integration, cell clustering and annotation methodology for the Yale, MGH, and Stanford Post-ICI cohorts

For the Yale and Stanford cohorts, individual sample count matrices were loaded into Seurat v5 R package for quality control and log normalization (17). To exclude low-quality reads from cells with high mitochondrial gene expression, we removed cells with greater than 5% mitochondrial gene expression. Additionally, we removed cells for which less than 200 genes were detected. Samples from the Stanford cohort were preprocessed before quality control to demultiplex the multiplexed samples. Demultiplexing was performed using Seurat using the published HTO tags with CLR normalization method. For the MGH cohort (16), we utilized the available count matrix from the published Seurat object.

Filtered samples from the Yale and Stanford cohorts were combined with MGH samples, batch corrected and integrated into the shared latent space using the scvi tool (18). We employed a grid search approach to optimize the integration from the three cohorts (number of latent dimensions = 10, training epochs = 3500, iLISI score = 0.443). This was followed by optimization of clustering using the shared scvi latent space with scanpy (19) using the neighborhood graph clustering algorithm with resolutions of 1 (number of neighbors = 50) for MGH, 1 (number of neighbors = 40) for Yale, and 0.8 (number of neighbors = 30) for Stanford, respectively. We employed the ScType unsupervised learning approach utilizing the comprehensive database of canonical gene markers to annotate the cell clusters (20).

**Supplemental Methods Text 8: Pathway Enrichment Characterization methodology**

We employed AUCell and VISION to calculate the gene-set enrichment scores for the gene modules associated with JAK-STAT, cytokine interaction, and pro-inflammatory immune response pathways such as NF-kB, TNF, Lipid and Atherosclerosis **(Supplemental Table 6)** (21,22). AUCell was used to first build the cell rankings of each gene in the gene expression dataset, which was used to calculate the AUC score of each gene signature in each cell. The identified active cells were then used as input to the VISION tool to calculate the autocorrelation scores (scale of 0-1) associating each gene set signature with the annotated cell types. Furthermore, we analyzed the graph structure of cytokine-cytokine interaction signaling using Cytoscape (23) and Protein-Protein Interaction (PPI) data from String database (24) (**Supplemental Figure 10**). The IL13 receptor emerged as one of the highly connected nodes. Pseudobulk expression analysis of 1L13 gene expression differences across myocarditis and control/nonmyocarditis samples within the Yale cohort revealed a significant difference in 1L13 expression (myocarditis samples = 0.012, control samples = 0.048, p-value = 0.03). These findings align with IL13 having the highest predictive value for myocarditis outcomes.

**Supplemental Table 6:** Table listing the set of genes used for characterizing cellular pathways in ICI-myocarditis using scRNA-seq data

| Pathways | Gene list |
| --- | --- |
| JAK_STAT_signaling_pathway | IL10, "IL21", "IL3", "IL6", "IL20", "TSLP", "IL15", "IL13", "IL9", "LIF", "IL2", "IL4", "IL5", "IL7", "IL11", "IL12A", "IL12B", "IL17A", "IL17B", "IL17C", "IL17D", "IL17E", "IL17F", "IL18", "IL19", "IL22", "IL23A", "IL24", "IL25", "IL26", "IL27", "IL28A", "IL28B", "IL29", "IL31", "IL32", "IL33", "CSF1", "CSF2", "CSF3", "EPO","EPOR", "GH1", "GH2", "GHR", "IFNA1", "IFNA2", "IFNA4", "IFNA5", "IFNA6", "IFNA7", "IFNA8", "IFNA10", "IFNA13", "IFNA14", "IFNA16", "IFNA17", "IFNA21", "IFNB1", "IFNG","IFNGR1", "IFNGR2", "IFNAR1", "IFNAR2", "IL10RA", "IL10RB", "IL11RA", "IL12RB1", "IL12RB2", "IL13RA1", "IL13RA2", "IL15RA", "IL17RA", "IL17RB", "IL17RC", "IL17RD", "IL17RE", "IL17RF", "IL18R1", "IL18RAP", "IL20RA", "IL20RB", "IL21R", "IL22RA1", "IL22RA2", "IL23R", "IL27RA", "IL28RA", "IL31RA", "CSF1R", "CSF2RA", "CSF2RB", "CSF3R", "JAK1", "JAK2", "JAK3", "TYK2", "STAT1", "STAT2", "STAT3", "STAT4", "STAT5A", "STAT5B", "STAT6", "PIK3CA", "PIK3CB", "PIK3CD", "PIK3CG", "AKT1", "AKT2", "AKT3", "MTOR", "PTPN1", "PTPN2", "PTPN6", "PTPN11", "SOCS1", "SOCS2", "SOCS3", "SOCS4", "SOCS5", "SOCS6", "SOCS7", "CISH", "GRB2", "SHC1", "IRS1", "IRS2", "MAPK1", "MAPK3", "MAPK8", "MAPK9", "MAPK10", "NFKB1", "RELA", "FOXO1", "FOXO3", "FOXO4" |
| JAK1_STAT1_pathway | "IFNAR1, "IFNAR2", "IFNG", "JAK1", "JAK2", "TYK2", "STAT1" |
| JAK1_STAT3_pathway | "IL6, "IL10", "IL11", "OSM", "JAK1", "JAK2", "STAT3" |
| JAK2_STAT5_pathway | "EPO, "TPO", "PRL", "GH1", "JAK2", "STAT5A", "STAT5B" |
| JAK3_STAT5_pathway | "IL2, "IL4", "IL7", "IL9", "IL15", "IL21", "JAK3", "STAT5A", "STAT5B" |
| TYK2_STAT4_pathway | "IL12A, "IL12B","IFNAR1", "IFNAR2", "TYK2", "JAK2", "STAT4" |
| JAK1_TYK2_STAT2_pathway | "IFNAR1, "IFNAR2", "JAK1", "TYK2", "STAT1", "STAT2", "STAT4" |
| JAK1_JAK2_STAT6_pathway | "IL4", "IL13", "JAK1", "JAK2", "STAT6" |
| Cytokine_cytokine_receptor_interaction | "CCL13", "CXCL10", "IL5", "IL15", "IL13", "TNFSF10", "CCL2", "IL12B", "IL27", "CCL17", "IL1A", "IL1B", "IL2", "IL3", "IL4", "IL6", "IL7", "IL9", "IL10", "IL11", "IL12A", "IL17A", "IL17B", "IL17C", "IL17D", "IL17E", "IL17F", "IL18", "IL19", "IL20", "IL21", "IL22", "IL23A", "IL24", "IL25", "IL26", "IL28A", "IL28B", "IL29", "IL31", "IL32", "IL33", "CSF1", "CSF2", "CSF3", "CCL1", "CCL3", "CCL4", "CCL5", "CCL7", "CCL8", "CCL11", "CCL14", "CCL15", "CCL16", "CCL18", "CCL19", "CCL20", "CCL21", "CCL22", "CCL23", "CCL24", "CCL25", "CCL26", "CCL27", "CCL28", "CXCL1", "CXCL2", "CXCL3","CXCL5", "CXCL6", "CXCL8", "CXCL9", "CXCL11", "CXCL12", "CXCL13", "CXCL14", "CXCL16", "CXCL17", "XCL1", "XCL2", "TNF", "LTA", "LTB", "FASLG", "CD70", "CD40LG", "TNFSF8", "TNFSF9", "TNFSF11", "TNFSF12", "TNFSF13", "TNFSF13B", "TNFSF14", "TNFSF15", "TNFSF18", "TNFSF4", "TNFSF5", "TNFSF6", "TNFSF7" |
| Allograft_rejection_pathway | "CD55", "IL17A", "TGFB1", "CD40", "GZMB", "CD28", "STAT1", "CASP8", "CTLA4", "PECR", "IL10", "VIM", "COL5A1", "IL2RA", "IL1A", "IL1B", "IL2", "IL21", "IL12A", "C8B", "C1QC", "PRKCZ", "CCL19", "C8A", "C1QB", "C1QA", "CASP9", "PDGFRA", "CXCL9", "CXCL11", "CXCL13", "CXCL12", "CASP3" , "CSNK2A2" , "CXCR5" , "FOXP3" , "CD40LG", "PRF1", "IL22", "IFNG", "IL12B", "IL13", "IL4", "IL5", "BHMT2", "C6", "C9", "LRRK2", "GDNF", "TUBA1B", "CD86", "CD80", "GABPA", "C4B", "C4A", "C2", "TNF", "VEGFA", "C3", "CASP7", "FAS", "MICA", "CCL21", "ABCB1", "C7", "HLA-DOA", "HLA-F", "HLA-DQA1", "C5", "AGTR1", "GNLY", "HLA-DMB", "HLA-DRA", "FASLG", "HLA-DPB1" ,"HLA-DMA", "HLA-DPA1", "HLA-G", "HLA-A", "HLA-DRB5", "HLA-E", "HLA-DOB" , "HLA-B" , "HLA-C" , "HLA-DQB1", "HLA-DQA2", "HLA-DRB1", "CXCL8" , "LOC110384692" , "HARS1" |
| Graft_versus_host_disease | "GZMB, "CD28", "IL1A", "IL1B" , "IL2" , "IL6" , "KLRC1", "KLRD1", "KIR3DL2" , "KIR3DL1" , "PRF1" , "IFNG", "CD86", "CD80" , "TNF" , "FAS" , "HLA-DOA" , "HLA-F" , "KIR2DL1" , "KIR2DL3" , "HLA-DQA1" , "HLA-DMB" , "HLA-DRA" , "FASLG" , "HLA-DPB1", "HLA-DMA" , "HLA-DPA1" , "HLA-G" , "HLA-A" , "HLA-DRB5", "HLA-E" , "HLA-DOB" , "HLA-B" , "HLA-C" , "HLA-DQB1" , "KIR2DL5A" , "KIR2DL2" , "HLA-DQA2" , "HLA-DRB1" , "HLA-DRB3", "HLA-DRB4" |
| NF_kappa_B_signaling_pathway | "CXCL12, "CCL4", "CXCL1", "NFKB1", "RELA", "RELB", "NFKB2", "IKBKB", "IKBKG", "TNF", "IL1A", "IL1B", "IL6", "TLR2", "TLR4", "TLR5", "TLR9", "MYD88", "TRAF6", "CHUK" |
| TNF_signaling_pathway | "CXCL10, "IL15", "CCL2", "TNF", "TNFRSF1A", "TNFRSF1B", "TRADD", "TRAF2", "NFKB1", "RELA" |
| Toll_like_receptor_signaling_pathway | "CD86", "CCL5", "CCL4", "CCL3", "IFNAR1" |
| Lipid_and_atherosclerosis | "CD40LG, "CCL5", "IL1B", "TNFSF10", "CCL3", "CD14" |
| T_cell_receptor_signaling | "IL10, "IL2", "CD3E", "CD3D", "CD3G", "CD4", "CD8A", "CD28", "CTLA4", "LCK", "ZAP70", "LAT", "ITK", "GRAP2", "SH2D1A", "NFATC1", "NFATC2", "NFATC3", "NFATC4", "MAPK1", "MAPK3","MAPK8", "MAPK9", "MAPK10", "RAC1", "RAC2", "CDC42", "NFKB1", "RELA", "FOS", "JUN" |

**Supplemental Methods Text 9: MOFA data processing methodology**

Two datasets (1 single‑cell RNA-seq dataset Stanford Post-ICI and 1 flow cytometry dataset) were used for MOFA analysis to determine immune signaling patterns related to CHIP and myocarditis status. The scRNA‑seq dataset profiled the targeted panel of 55 cytokine genes. The Stanford scRNA-seq cohort (pre- and post-steroid samples) has seventeen non-myocarditis (Group A, B) and eight myocarditis samples (Group C) with 97,367 total cells; six samples carry CHIP (A5, A9, B3, B10, B13, MCL3) and 19 do not, the post-steroid samples are B2, B10, MCE7 and MCL3, and the group-A samples have no recorded steroid timing and are excluded from the steroid-stratified comparisons. Finally, Stanford CyTOF data on ICI-induced myocarditis were included, with 16 non-myocarditis samples (2 CHIP, 14 non-CHIP) and 6 myocarditis samples (1 CHIP, 5 non-CHIP).

The CyTOF data were preprocessed in R. Arcsine transformation and bead normalization were performed using the CATALYST package, followed by live-cell gating via the gate_flowclust_2d algorithm in the openCyto package (25). The MOFA+ package (26) was used to extract latent factors from the datasets. The input data consisted of cytokine expression of 55 genes across the Stanford scRNA-seq data and channel intensities from the Stanford CyTOF data. The samples were grouped by CHIP status within the myocarditis group (CHIP versus non-CHIP), myocarditis status (myocarditis versus non-myocarditis), and steroid timing (pre-steroid versus all samples (pre-steroid and post-steroid)). A separate MOFA model was fit to each group. MOFA analysis was implemented using the mofapy2 package in Python. After inspection of several trial runs, K=15 latent factors were used, with a spike-and-slab prior and group-wise ARD priors for automatic relevance determination to ensure feature weights were sparse. Stanford count data were normalized per cell within the 55-cytokine panel (counts per 10,000) and log1p-transformed. Models were trained for up to 2000 iterations to medium convergence. After training, the top 20 features (cytokines) explaining most of the variance were extracted and compared across CHIP vs. non-CHIP within myocarditis samples, myocarditis vs. non-myocarditis samples, and pre-steroid vs. all samples (pre steroid and post-steroid). The Enrichr tool (27) was used to extract KEGG pathway information from the set of top features for the myocarditis-stratified and steroid-stratified models. The Stanford scRNA and CyTOF data were additionally integrated by a joint MOFA over scRNA cytokine expression and cell-type-resolved CyTOF marker means.

**Supplemental Table 7:** Table showing number of CHIP mutations among patients on ICI stratified by myocarditis outcome

|  | No myocarditis | myocarditis | P |
| --- | --- | --- | --- |
| Total | 38 (43%) | 50 (57%) | 0.003 |
| #CHIP mutations |  |  |  |
| 0 | 28 (74%) | 17 (34%) |  |
| 1 | 8 (21%) | 25 (50%) |  |
| 2 | 1 (2.6%) | 5 (10%) |  |
| 3 | 1 (2.6%) | 3 (6%) |  |

**Supplemental Figure 1:** Bar graph showing distribution of entire cohort by number of CHIP mutations

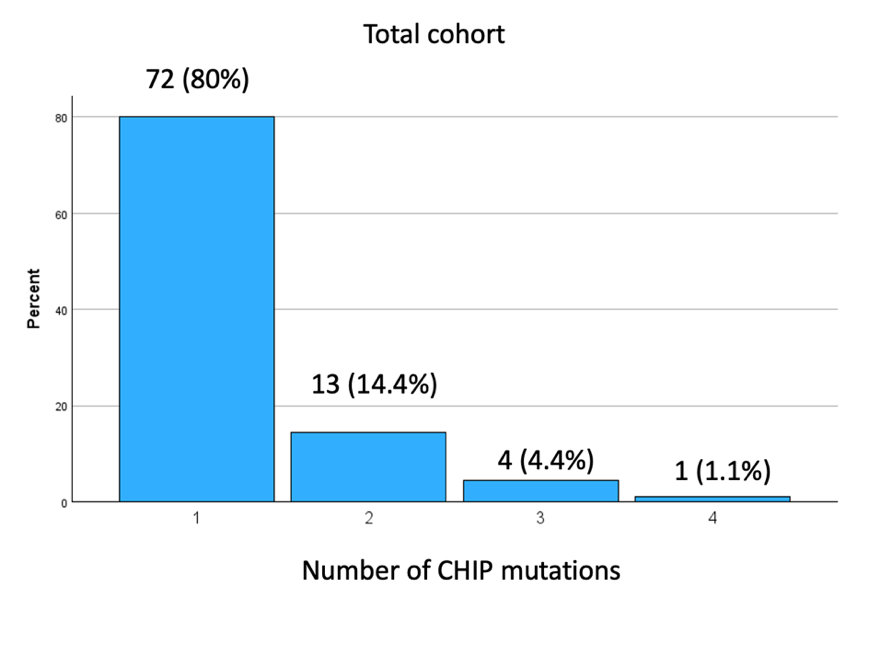

**Supplemental Table 8:** Table of CHIP Variant Frequencies a) Among all CHIP patients* b) Among CHIP Patients on ICI with myocarditis** c) Among CHIP Patients on ICI who did not develop myocarditis***.

*Frequency of RUNX1 0%

**Frequency of IDH2, ZRSR2, KMT2B, NRAS, RUNX1, SRSF2, TET1, CREBBP, FBXW7, FLT2, BCL11B is 0%

***Frequency of CREBP, TP53, FLT3, KDM6A, CBL, NRAS, RUNX1, SRSF2, TET1, U2AF1, SF3B1, ZRSR2, NOTCH1, CSF3R, JAK2 is 0%

A)

| **CHIP gene** | DNMT3A | TET2 | PPM1D | EZH2 | TP53 | IDH2 | ASXL1 | JAK2 | SRSF2 | CBL | SF3B1 | CREBBP | CSF3R | TET1 | U2AF1 | ZRSR2 | NOTCH1 | NRAS | FBXW7 | FLT3 | KDM6A | KMT2B | BCL11B |
| --- | --- | --- | --- | --- | --- | --- | --- | --- | --- | --- | --- | --- | --- | --- | --- | --- | --- | --- | --- | --- | --- | --- | --- |
| Frequency | 33.00 | 13.00 | 10.00 | 10.00 | 8.00 | 7.00 | 5.00 | 5.00 | 4.00 | 3.00 | 2.00 | 2.00 | 2.00 | 1.00 | 1.00 | 1.00 | 1.00 | 1.00 | 1.00 | 1.00 | 1.00 | 1.00 | 1.00 |
| Percentage | 35.87 | 14.13 | 10.87 | 10.87 | 8.70 | 7.61 | 5.43 | 5.43 | 4.35 | 3.26 | 2.17 | 2.17 | 2.17 | 1.09 | 1.09 | 1.09 | 1.09 | 1.09 | 1.09 | 1.09 | 1.09 | 1.09 | 1.09 |

B)

| **CHIP gene** | DNMT3A | TET2 | PPM1D | EZH2 | TP53 | ASXL1 | CBL | JAK2 | CSF3R | KDM6A | U2AF1 | SF3B1 | NOTCH1 |
| --- | --- | --- | --- | --- | --- | --- | --- | --- | --- | --- | --- | --- | --- |
| Frequency | 12 | 8 | 6 | 3 | 3 | 3 | 3 | 2 | 1 | 1 | 1 | 1 | 1 |
| Percentage | 34.29 | 22.86 | 17.14 | 8.57 | 8.57 | 8.57 | 8.57 | 5.71 | 2.86 | 2.86 | 2.86 | 2.86 | 2.86 |

C)

| **CHIP gene** | DNMT3A | ASXL1 | PPM1D | EZH2 | TET2 | IDH2 | FBXW7 | BCL11B | KMT2B |
| --- | --- | --- | --- | --- | --- | --- | --- | --- | --- |
| Frequency | 3 | 2 | 2 | 1 | 1 | 1 | 1 | 1 | 1 |
| Percentage | 30 | 20 | 20 | 10 | 10 | 10 | 10 | 10 | 10 |

**Supplemental Table 9:** Cancer Subtypes Among ICI recipients stratified by CHIP Status

| Cancer type | No Chip | Chip | total | pvalue |
| --- | --- | --- | --- | --- |
| Breast | 15.2%(7/46) | 9.5%(4/42) | 12.5%(11/88) | 0.526 |
| GI | 4.3%(2/46) | 4.8%(2/42) | 4.5%(4/88) | 1.000 |
| GU | 21.7%(10/46) | 28.6%(12/42) | 25%(22/88) | 0.473 |
| Gyn | 2.2%(1/46) | 2.4%(1/42) | 2.3%(2/88) | 1.000 |
| H&N | 2.2%(1/46) | 4.8%(2/42) | 3.4%(3/88) | 0.604 |
| Liver | 4.3%(2/46) | 2.4%(1/42) | 3.4%(3/88) | 1.000 |
| Lung | 28.3%(13/46) | 23.8%(10/42) | 26.1%(23/88) | 0.809 |
| Melanoma | 15.2%(7/46) | 14.3%(6/42) | 14.8%(13/88) | 1.000 |
| Mesothelioma | 2.2%(1/46) | 2.4%(1/42) | 2.3%(2/88) | 1.000 |
| Sarcoma | 0%(0/46) | 4.8%(2/42) | 2.3%(2/88) | 0.225 |
| Skin | 4.3%(2/46) | 2.4%(1/42) | 3.4%(3/88) | 1.000 |

**Supplemental Table 10:** Demographics and Clinical Characteristics of 12 MSKCC Patients and 6 Non-Cancer Controls

|  | Demographics | | Cancer Treatment | | Comorbidities | | | | |
| --- | --- | --- | --- | --- | --- | --- | --- | --- | --- |
|  | Gender (%F) | Mean Age (SD) | ICI | XRT  (Chest) | CHIP | HFrEF | DM | CAD/  CAC | CM |
| 12 breast cancer  patients  (MSKCC) | 100%  (12/12) | 66.6  (12.3) | 8.3% (1/12) | 41.6%  (5/12) | 100% (12/12) | 0% | 25%(  3/12) | 8.3%  (1/12) | 0% |
| 6 non-cancer patients | 50%  (3/6) | 55.2  (13.6) | NA | NA | 33.3% (2/6) | 0% | 33.3%(  2/6) | 16.7%  (1/6) | 0% |

**Supplemental Figure 2:** Correlation plots of individual cytokines with myocarditis

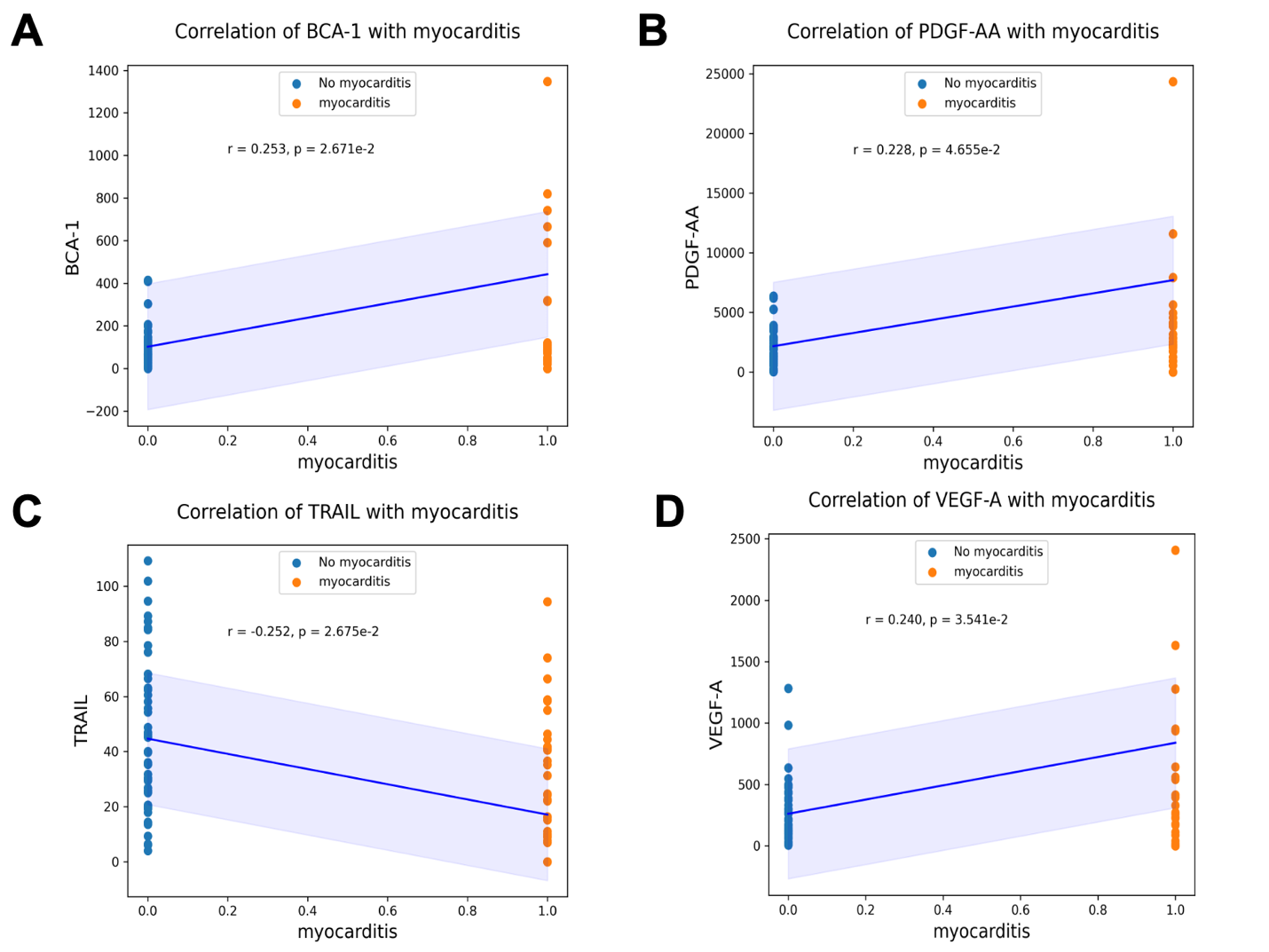

**Supplemental Figure 3:** Correlation plots of individual cytokines with immunosuppression resistant myocarditis

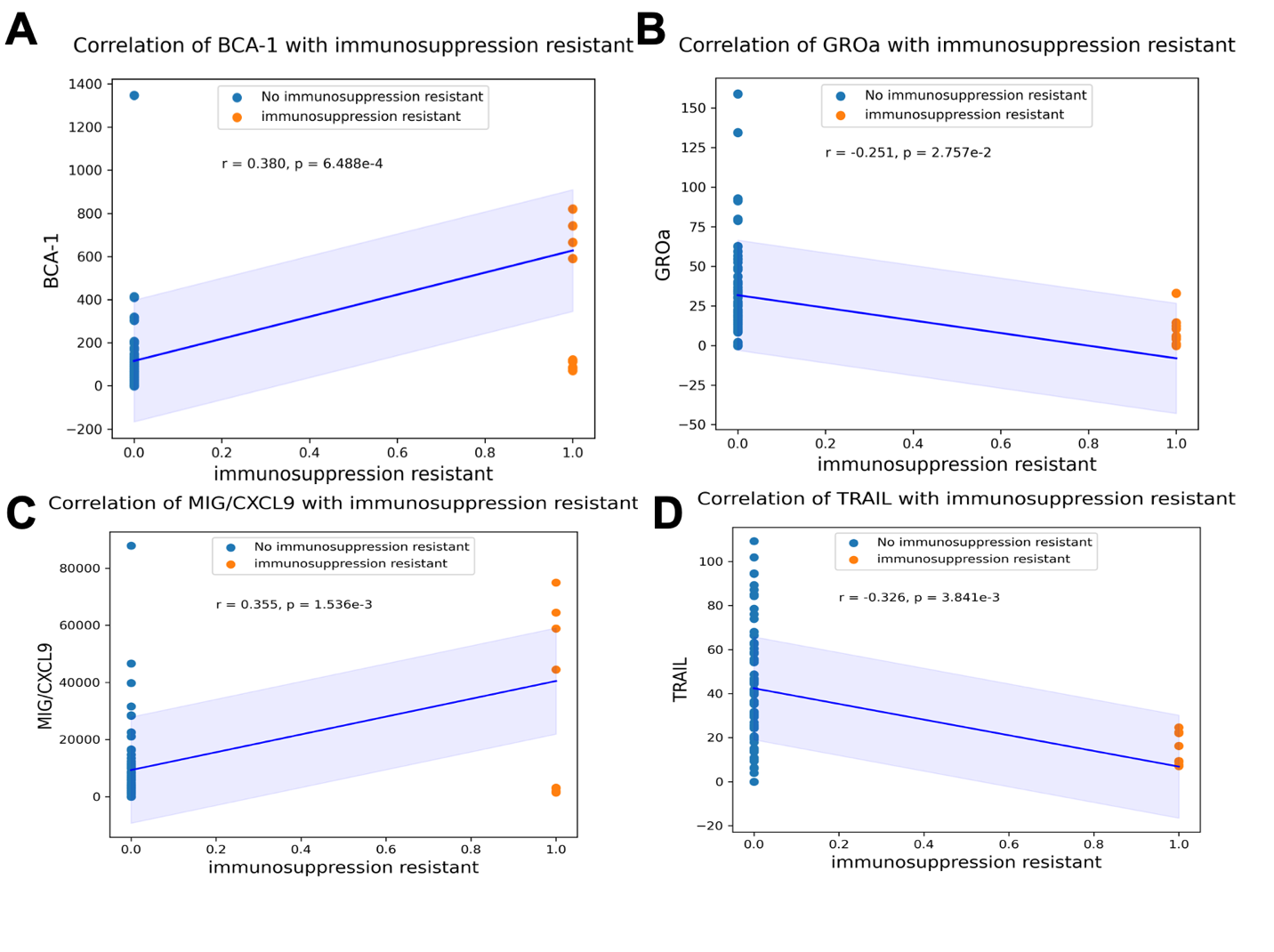

**Supplemental Table 11:** Cytokines identified by XGBoost and Corresponding Gene Names

| **Cytokine in myocarditis model** | **Gene Name** | **Cytokine (Immuno-suppression resistant)** | **Gene Names** |
| --- | --- | --- | --- |
| IL13 | IL13 | MIP1b | CCL4 |
| IP10 | CXCL10 | IL10 | IL10 |
| PDGF-AA | PDGFA | Eotaxin 2 | CCL24 |
| TRAIL | TNFSF10 | MDC | CCL22 |
| IL5 | IL5 | TNFb | TNFB |
| MCP-1 | CCL2 | ENA-78 | CXCL5 |
| MCP-4 | CCL13 | IL1a | IL1A |
| TARC | CCL17 | BCA1 | CXCL13 |
| SCF | KITLG | FGF2 | FGF2 |
| FGF2 | fgf2 | EGF | EGF |
| IL12P40 | IL12B | GROa | CXCL1 |
| IL15 | IL15 | SDF1 a+b | CXCL12, CXCL12b |
| IL27 | IL27 |  |  |

**Supplemental Table 12:** All significant pathways for myocarditis model

| term | p-value | q-value |
| --- | --- | --- |
| Cytokine-cytokine receptor interaction | 0.000 | 0.000 |
| IL-17 signaling pathway | 0.000 | 0.000 |
| Viral protein interaction with cytokine and cytokine receptor | 0.000 | 0.000 |
| Pathways in cancer | 0.000 | 0.000 |
| JAK-STAT signaling pathway | 0.000 | 0.000 |
| Influenza A | 0.000 | 0.000 |
| Chemokine signaling pathway | 0.000 | 0.000 |
| Inflammatory bowel disease | 0.000 | 0.000 |
| Th1 and Th2 cell differentiation | 0.000 | 0.000 |
| TNF signaling pathway | 0.000 | 0.000 |
| Asthma | 0.000 | 0.001 |
| Allograft rejection | 0.000 | 0.002 |
| Rap1 signaling pathway | 0.000 | 0.002 |
| Lipid and atherosclerosis | 0.000 | 0.002 |
| Coronavirus disease | 0.000 | 0.002 |
| Ras signaling pathway | 0.000 | 0.002 |
| Intestinal immune network for IgA production | 0.000 | 0.002 |
| MAPK signaling pathway | 0.001 | 0.003 |
| Fc epsilon RI signaling pathway | 0.001 | 0.003 |
| RIG-I-like receptor signaling pathway | 0.001 | 0.003 |
| Melanoma | 0.001 | 0.003 |
| PI3K-Akt signaling pathway | 0.001 | 0.005 |
| Rheumatoid arthritis | 0.002 | 0.005 |
| Hematopoietic cell lineage | 0.002 | 0.005 |
| Chagas disease | 0.002 | 0.005 |
| C-type lectin receptor signaling pathway | 0.002 | 0.005 |
| Toll-like receptor signaling pathway | 0.002 | 0.005 |
| Fluid shear stress and atherosclerosis | 0.004 | 0.009 |
| Phospholipase D signaling pathway | 0.004 | 0.010 |
| Proteoglycans in cancer | 0.008 | 0.018 |
| Regulation of actin cytoskeleton | 0.009 | 0.020 |
| Calcium signaling pathway | 0.010 | 0.023 |
| African trypanosomiasis | 0.024 | 0.053 |
| Type I diabetes mellitus | 0.028 | 0.059 |
| Malaria | 0.032 | 0.067 |
| Autoimmune thyroid disease | 0.034 | 0.069 |
| Legionellosis | 0.036 | 0.072 |
| Cytosolic DNA-sensing pathway | 0.040 | 0.075 |
| Herpes simplex virus 1 infection | 0.040 | 0.075 |
| Glioma | 0.048 | 0.085 |
| Pertussis | 0.048 | 0.085 |
| Leishmaniasis | 0.049 | 0.085 |

**Supplemental Table 13:** All significant pathways for immunosuppression resistant myocarditis model

| term | p-value | q-value |
| --- | --- | --- |
| Viral protein interaction with cytokine and cytokine receptor | 0.000 | 0.000 |
| Cytokine-cytokine receptor interaction | 0.000 | 0.000 |
| Chemokine signaling pathway | 0.000 | 0.000 |
| Rheumatoid arthritis | 0.000 | 0.000 |
| Pertussis | 0.000 | 0.000 |
| NF-kappa B signaling pathway | 0.000 | 0.001 |
| Regulation of actin cytoskeleton | 0.000 | 0.004 |
| Intestinal immune network for IgA production | 0.000 | 0.004 |
| Inflammatory bowel disease | 0.001 | 0.006 |
| MAPK signaling pathway | 0.001 | 0.006 |
| Melanoma | 0.001 | 0.007 |
| Leishmaniasis | 0.001 | 0.007 |
| IL-17 signaling pathway | 0.002 | 0.010 |
| Amoebiasis | 0.002 | 0.011 |
| C-type lectin receptor signaling pathway | 0.002 | 0.011 |
| TNF signaling pathway | 0.002 | 0.012 |
| FoxO signaling pathway | 0.003 | 0.015 |
| Breast cancer | 0.004 | 0.017 |
| Gastric cancer | 0.004 | 0.017 |
| Pathways in cancer | 0.004 | 0.017 |
| JAK-STAT signaling pathway | 0.005 | 0.018 |
| Tuberculosis | 0.006 | 0.021 |
| Kaposi sarcoma-associated herpesvirus infection | 0.007 | 0.023 |
| Rap1 signaling pathway | 0.008 | 0.026 |
| Human cytomegalovirus infection | 0.009 | 0.029 |
| Ras signaling pathway | 0.010 | 0.029 |
| Chemical carcinogenesis | 0.010 | 0.029 |
| Calcium signaling pathway | 0.010 | 0.029 |
| Asthma | 0.020 | 0.055 |
| PI3K-Akt signaling pathway | 0.021 | 0.057 |
| African trypanosomiasis | 0.024 | 0.061 |
| Allograft rejection | 0.024 | 0.061 |
| Bladder cancer | 0.026 | 0.063 |
| Graft-versus-host disease | 0.027 | 0.063 |
| Type I diabetes mellitus | 0.028 | 0.063 |
| Malaria | 0.032 | 0.071 |
| Autoimmune thyroid disease | 0.034 | 0.073 |
| Legionellosis | 0.036 | 0.076 |
| Endometrial cancer | 0.037 | 0.076 |
| Cytosolic DNA-sensing pathway | 0.040 | 0.080 |
| Epithelial cell signaling in Helicobacter pylori infection | 0.045 | 0.087 |
| Non-small cell lung cancer | 0.046 | 0.087 |
| Glioma | 0.048 | 0.088 |
| Pancreatic cancer | 0.048 | 0.088 |

**Supplemental Table 14:** Cytokines significantly associated with high risk CHIP score by point biserial correlation

| Cytokine | R value | P-Value |
| --- | --- | --- |
| FGF-2 | 0.31 | 0.003 |
| IL-6 | 0.28 | 0.01 |
| IL-8 | 0.28 | 0.01 |
| IL-5 | 0.28 | 0.01 |

**Supplemental Figure 4:** SHAP model (left) and graph of top implicated KEGG pathways (right) for top cytokine features associated with high and intermediate risk CHIP

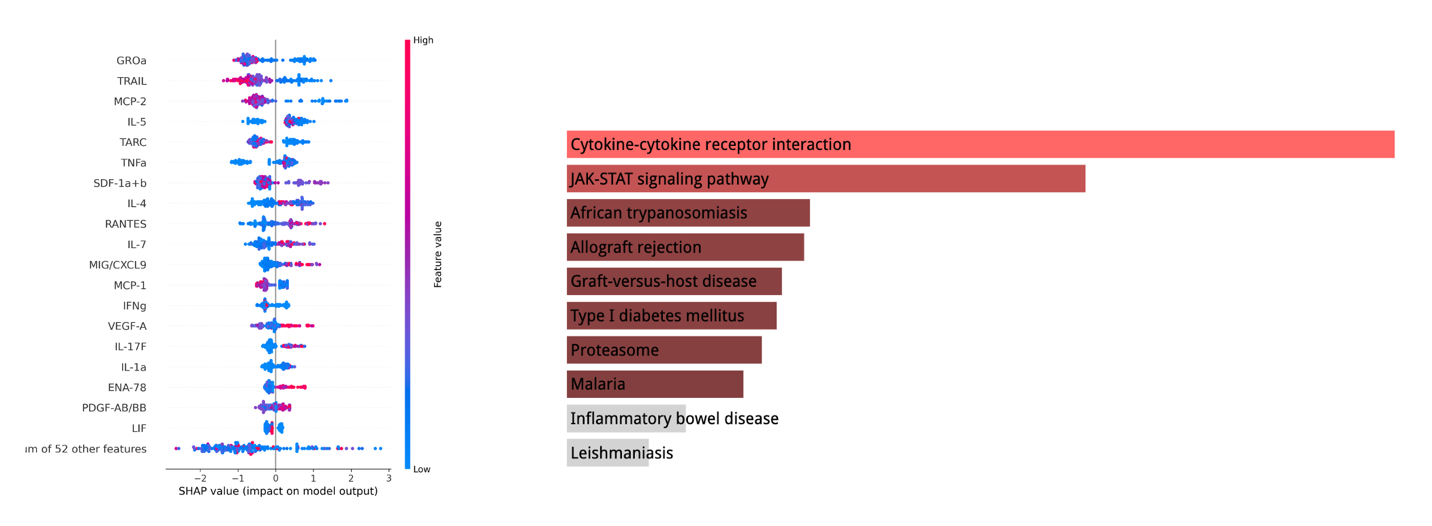

**Supplemental Figure 5:** Graph of top 10 implicated KEGG pathways associated with both ICI myocarditis (A) and intermediate to high risk CHIP (B) using cytokine features from the Stanford validation cohort .

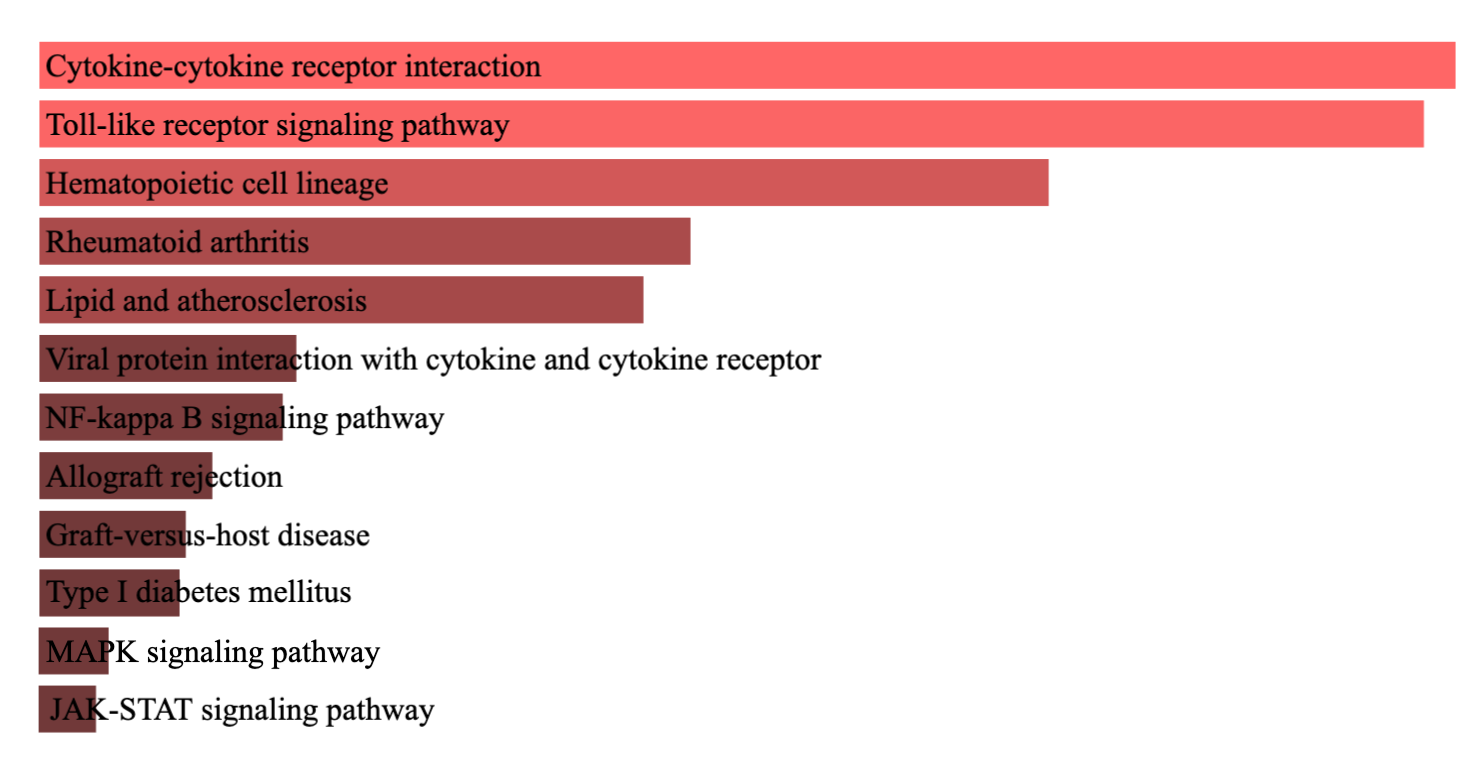

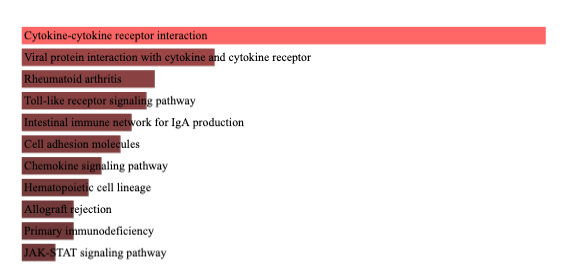

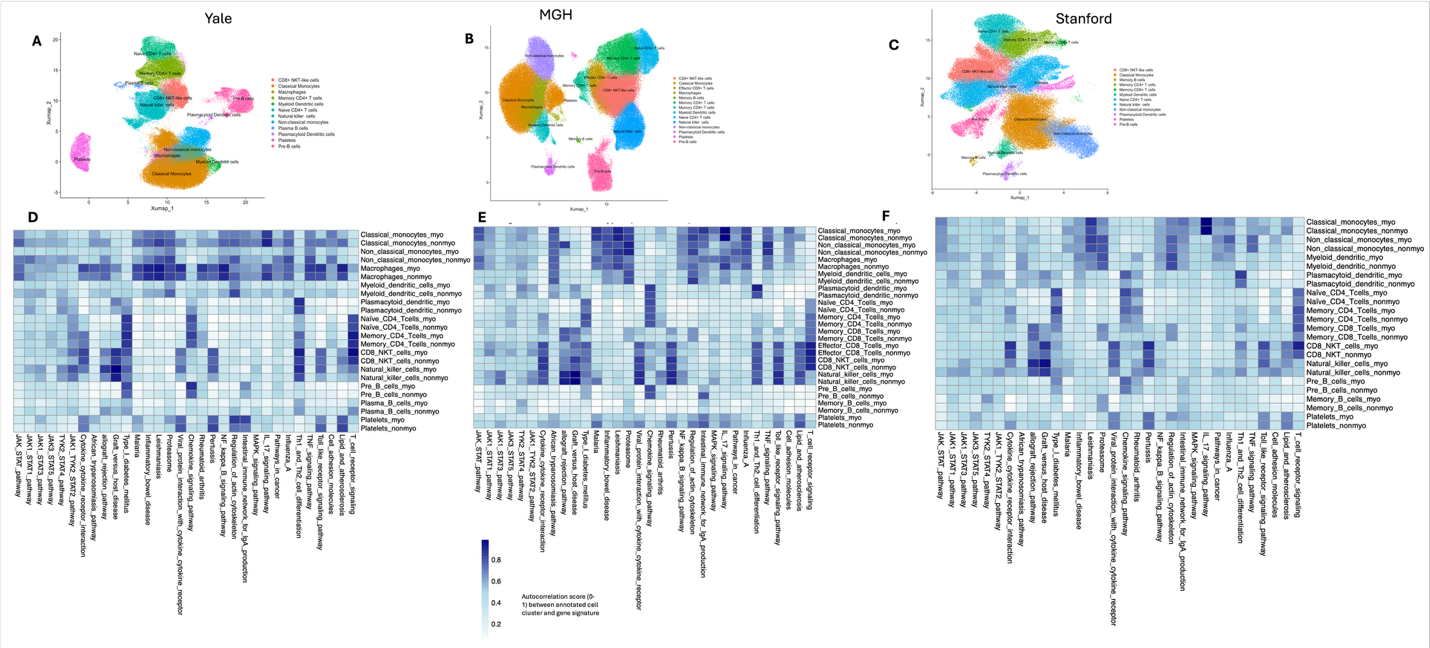

**Supplemental Figure 6: A, B,C** Umap of annotated cell populations in the MGH, YALE, and Stanford cohorts. **C, D**, **E** Heatmap illustrating differences in the autocorrelation scores between Myocarditis and Non-myocarditis (control) cells, demonstrating the cell activity of gene programs in these two conditions for all PBMC scRNA-seq samples (pre-steroid and post-steroid samples) from MGH, YALE, and Stanford cohorts.

| **PatientID** | **CHIP**  **status** | **Steroid**  **status** | **Sex** | **Age** | **Cancer_type** | **ICI_agent** | **Myocarditis status** |
| --- | --- | --- | --- | --- | --- | --- | --- |
| 1 | CHIP | Pre | F | 71-80 | lung | durvalumab | Myocarditis |
| 2 | CHIP | Pre | F | 71-80 | melanoma | nivolumab | Myocarditis |
| 3 | CHIP | Pre | F | 61-70 | lung | durvalumab | Myocarditis |
| 4 | CHIP | Pre | M | 71-80 | lung | pembrolizumab | Myocarditis |
| 5 | CHIP | Pre | M | 61-70 | melanoma | Nivolumab/ ipilimumab | Myocarditis |
| 6 | CHIP | Pre | M | 71-80 | RCC | pembrolizumab | Myocarditis |
| 7 | CHIP | Pre | M | >80 | bladder | Pembrolizumab | Myocarditis |
| 8 | CHIP | **Post** | M | >80 | bladder | Pembrolizumab | Myocarditis |
| 9 | Non-CHIP | Pre | M | 71-80 | sarcoma | C1D1 Keytruda (pembro) | Myocarditis |
| 10 | Non-CHIP | Pre | M | >80 | melanoma | Pembrolizumab | Myocarditis |
| 11 | Non-CHIP | Pre | F | 61-70 | lung | Atezolizumab | Myocarditis |
| 12 | Non-CHIP | Pre | M | <50 | lung | Pembrolizumab | Myocarditis |
| 13 | Non-CHIP | **Post** | M | >80 | melanoma | Nivolumab/ ipilimumab | Myocarditis |
| 14 | CHIP | **Pre** | F | 51-60 | GU | pembrolizumab | Non-myocarditis |
| 15 | CHIP | **Pre** | M | 71-80 | bladder | None | Non-myocarditis |
| 16 | CHIP | **Pre** | M | 61-70 | lung | None | Non-myocarditis |
| 17 | Non-CHIP | **Pre** | M | 71-80 | melanoma | pembrolizumab | Non-myocarditis |
| 18 | Non-CHIP | **Pre** | F | 71-80 | lung | durvalumab | Non-myocarditis |
| 19 | Non-CHIP | **Pre** | F | 61-70 | Endomedtrial/GU | pembrolizumab | Non-myocarditis |

**Supplemental Table 15:** Demographics and Clinical Characteristics of 19 patients with PBMC tissue samples comprising internal Yale cohort in multiomics analysis

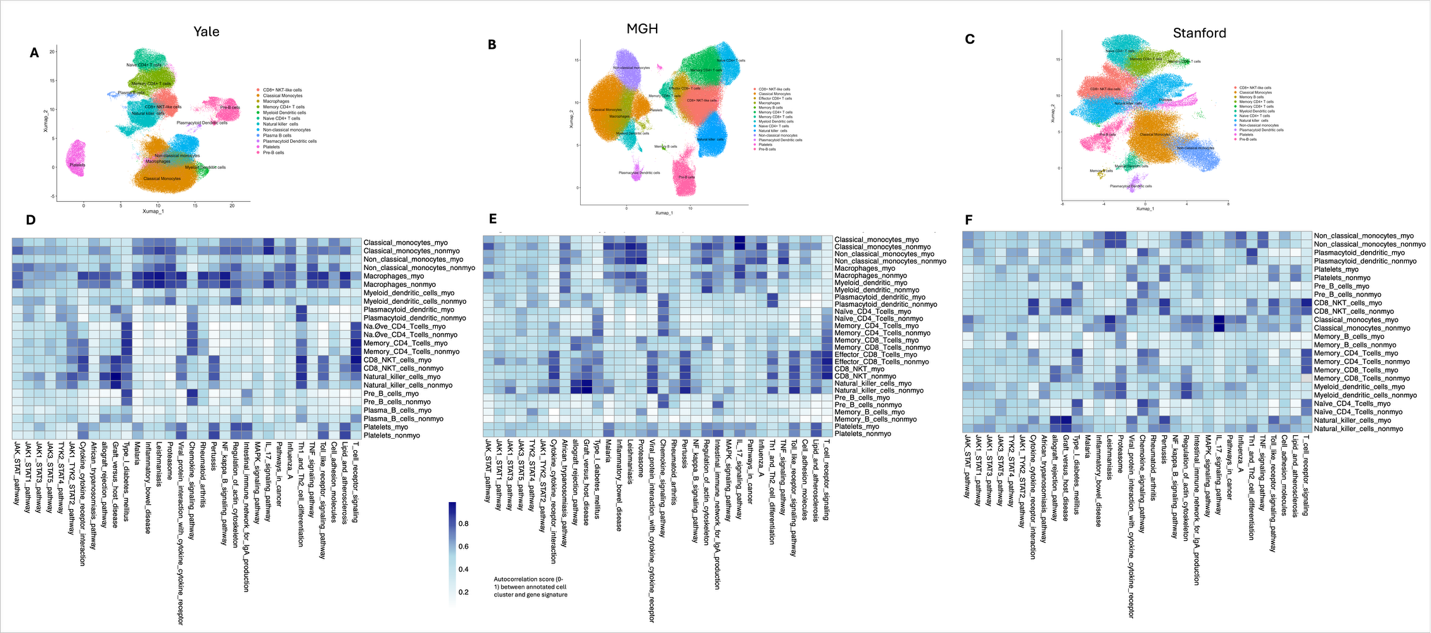

**Supplemental Figure 7:** **A, B,C** Umap of annotated cell populations in the MGH, YALE, and Stanford cohorts. **C, D**, **E** Heatmap illustrating differences in the autocorrelation scores between Myocarditis and Non-myocarditis (control) cells, demonstrating the cell activity of gene programs in these two conditions for pre-steroid PBMC scRNA-seq samples from MGH, YALE, and Stanford cohorts.

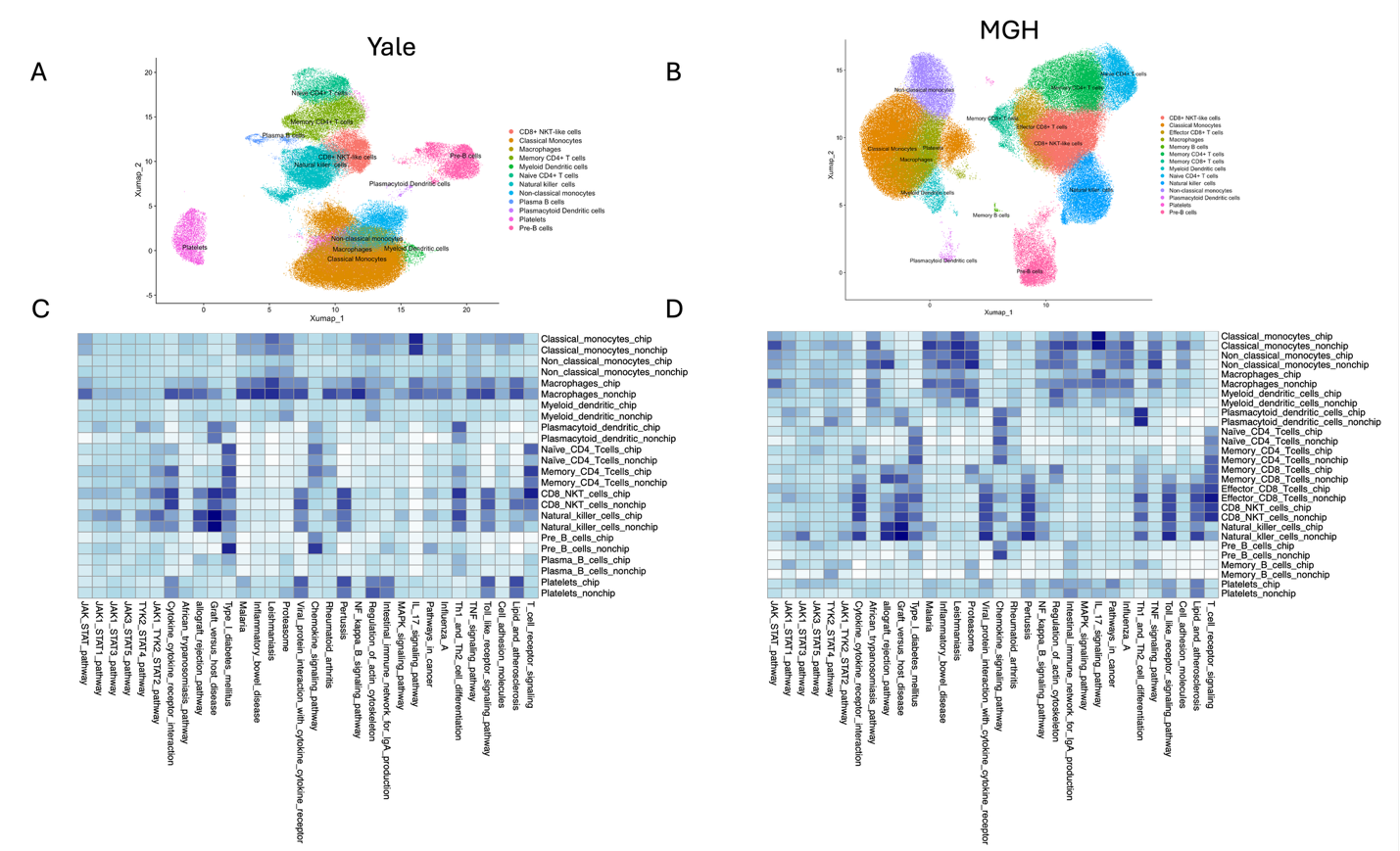

**Supplemental Figure 8:** **A, B** Umap of annotated cell populations in the MGH and YALE cohort. **C, D** Heatmap illustrating the differences in autocorrelation scores between Myocarditis CHIP and Myocarditis NONCHIP cells, demonstrating the cell activity of gene programs in these two conditions for all PBMC scRNA-seq samples (pre-steroid and post-steroid samples) from MGH and YALE cohorts.

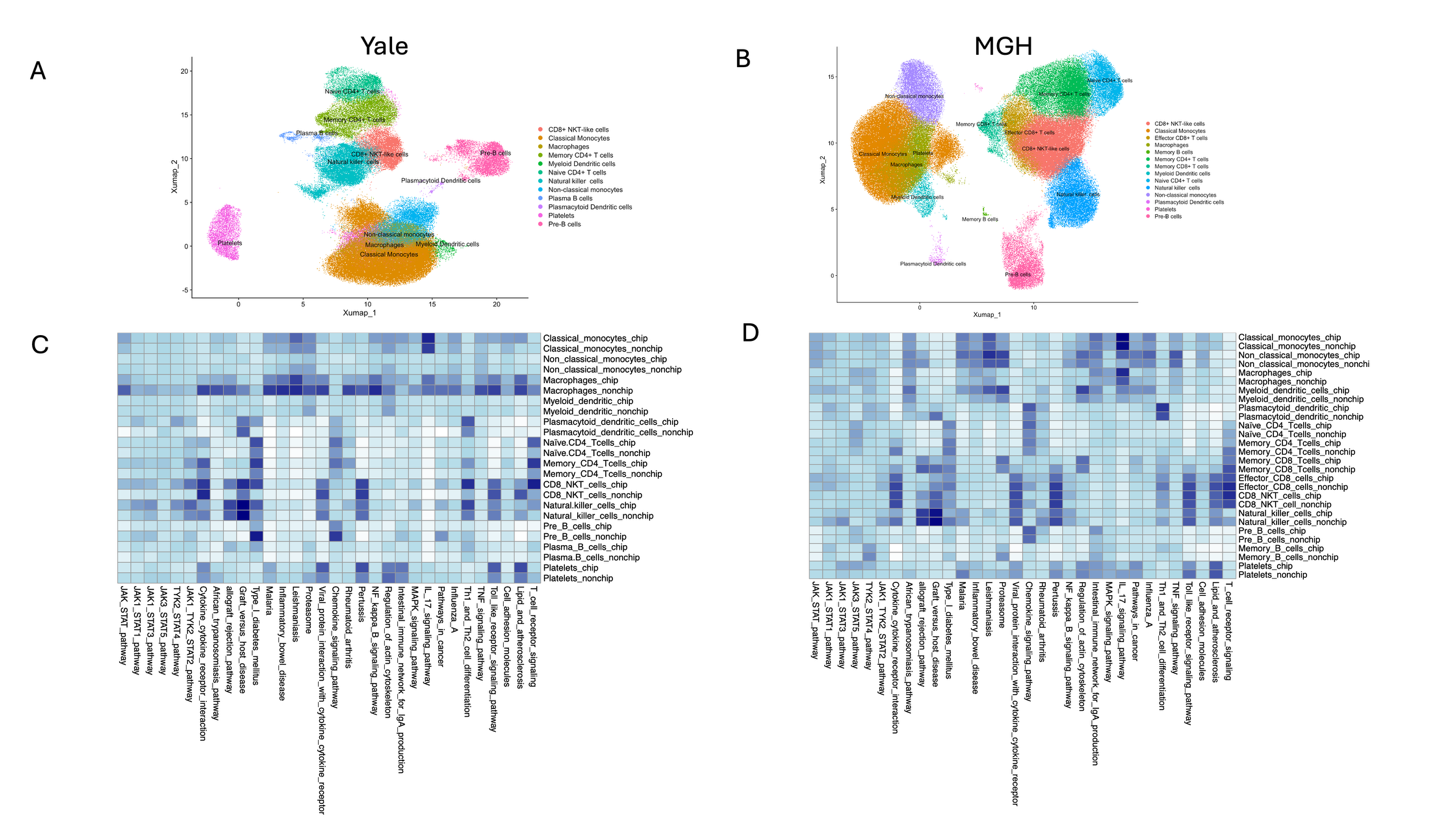

**Supplemental Figure 9:** Umap of annotated cell populations in the MGH and YALE cohort. **C, D** Heatmap illustrating the differences in autocorrelation scores between Myocarditis CHIP and Myocarditis NONCHIP cells, demonstrating the cell activity of gene programs in these two conditions for pre-steroid PBMC scRNA-seq samples from MGH and YALE cohorts.

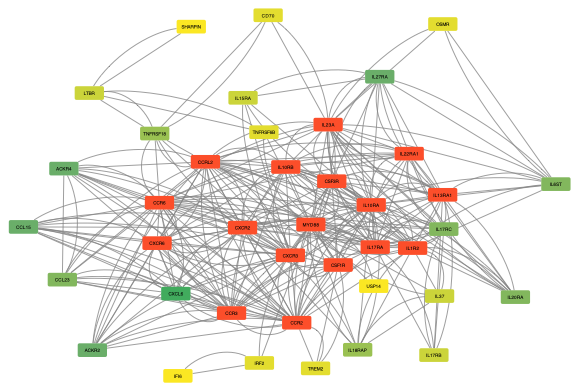
**Supplemental Figure 10:** Cytoscape visualization of the graph structure of cytokine-cytokine interaction signaling showing the high connectivity cytokines (represented by high degree nodes (red colored central nodes)).

**Supplemental Results Text 1** MOFA results

Comparing myocarditis and non-myocarditis samples in the Stanford cohort (**Supplemental Tables 16 and 17, Supplemental Figure 10**), the top variable cytokines include the myocardial chemokines CCL5, CCL4, and CCL3 and pro-inflammatory cytokines such as TNFSF10, IL1B, IL18, and CD40LG. Steroid timing has little effect on the dominant cytokine program: within the myocarditis group the pre-steroid and pre+post-steroid top-20 feature sets are identical, differing only in rank order, and within the non-myocarditis group they differ by a single feature, with CXCL8 among the top features before steroids and PDGFA after. Finally, integrating the scRNA and CyTOF data with a joint two-view MOFA recovers the three canonical immune axes (lymphoid, myeloid, and NK/cytotoxic) from the CyTOF markers, and with a cell-type-resolved CyTOF view surfaces a shared factor coupling pro-inflammatory cytokines (IL1B, TNF, CXCL8, IL23A, CCL2) to a coordinated T/NK differentiation pattern (exploratory, given the eleven patients profiled by both platforms).

##### Myocarditis-stratified

|  |  | **Stanford Cohort** |
| --- | --- | --- |
| **Myocarditis only** |  |  |
| **Both** |  | IL23A IFNG , CCL5 FLT3LG IFNAR1 TNFSF10 CSF3R IL1B CCL3 IL16 CCL4 IL15 CXCL8 CD40LG **PDGFA** IL18 LTA **TNF** IL2RA IL3RA |
| **Non-myocarditis only** |  | **VEGFA** |

**Supplemental Table 16:** Top features in the Stanford cohort, myocarditis-stratified.

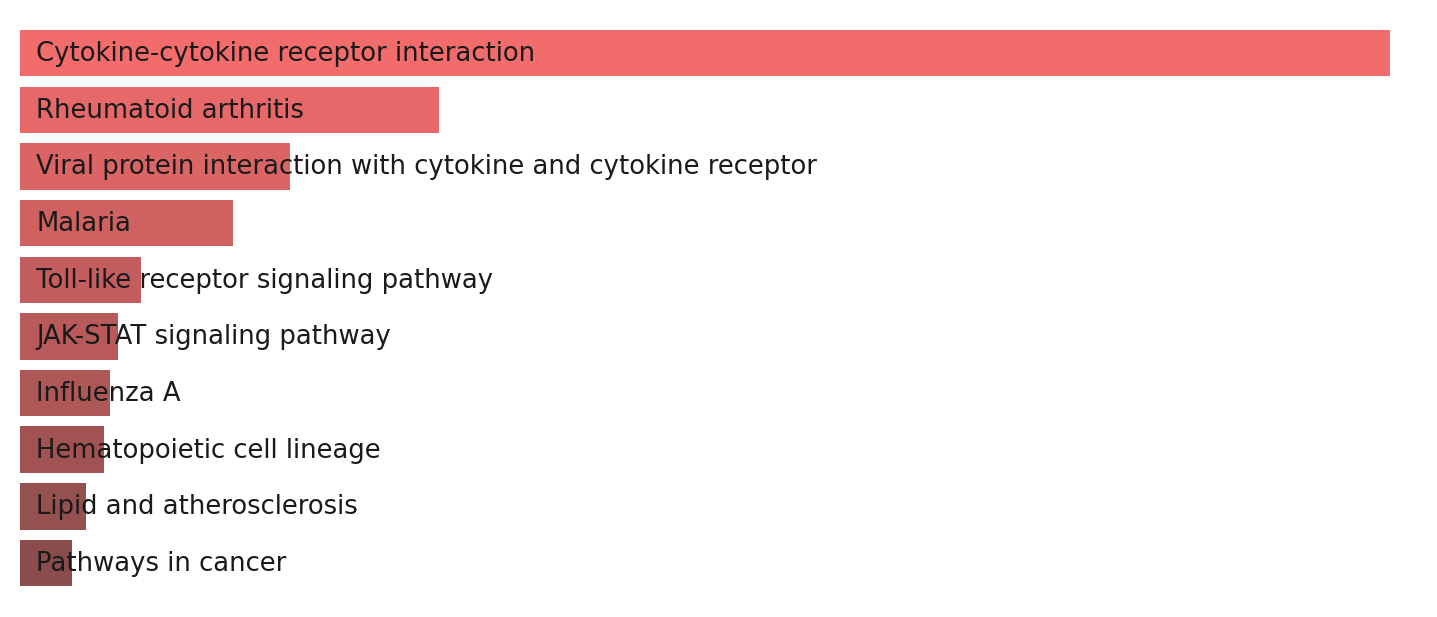

**Supplemental Figure 11:** Top KEGG pathways associated with the cytokines that differed between myocarditis and nonmyocarditis samples in the Stanford cohort (top features listed in Supplemental Table 11).

|  | **Pre-steroid only** | **Shared (pre and pre+post)** | **Pre+Post-steroid only** |
| --- | --- | --- | --- |
| **Myocarditis** | — | CCL5 TNFSF10 IFNAR1 IL16 CSF3R CCL4 FLT3LG IL18 CCL3 CD40LG IL1B TNF IL15 CXCL8 LTA IL3RA IL2RA IFNG IL23A PDGFA | — |
| **Non-myocarditis** | CXCL8 | IFNAR1 IL16 CCL5 FLT3LG TNFSF10 CSF3R CCL4 CD40LG IL15 IL18 CCL3 IL1B IL3RA LTA TNF IL2RA IL23A IFNG IL27 | PDGFA |

**Supplemental Table 17:** Pre-steroid versus pre+post-steroid top features for the Stanford cohort, stratified by myocarditis status. Stanford is the cohort with matched CyTOF and scRNA-seq data.
